# Cross-ancestry proteome-wide Mendelian randomization of plasma proteins and breast cancer susceptibility

**DOI:** 10.64898/2026.05.04.26352055

**Authors:** Xueyao Wu, Devika Godbole, Jacob Williams, Jayati Sharma, Jiyeon Choi, Zhonghua Liu, Peter Kraft, Haoyu Zhang

## Abstract

**Background:** The plasma proteome may help connect inherited breast cancer susceptibility to biologically interpretable and potentially actionable pathways, but proteome-wide evidence across ancestries remains limited.

**Methods:** We conducted proteome-wide Mendelian randomization (MR) analyses of breast cancer risk using *cis*-protein quantitative trait loci for 1,962 distinct plasma proteins from 34,557 European-ancestry participants in the UK Biobank Pharma Proteomics Project. Breast cancer genome-wide association studies included 156,901 cases and 204,634 controls of European, East Asian, and African ancestries. Ancestry-specific MR estimates were combined by fixed-effects meta-analysis. Candidate proteins were further evaluated through subtype-specific and reverse-direction MR analyses, statistical colocalization, replication in independent European and East Asian proteomic datasets, and analyses of associations between protein polygenic scores and breast cancer status in All of Us.

**Results:** Twelve proteins achieved Bonferroni significance in the cross-ancestry meta-analysis (*P* < 0.05/1,971 tests), including eight previously reported proteins and four newly prioritized by MR. DNPH1 showed cross-ancestry heterogeneity, while CASP8, RALB, and USP28 showed cross-subtype heterogeneity after accounting for shared controls (all *P* < 0.05/12). Complementary support varied: six candidates showed strong colocalization (PPH4 ≥ 0.80); four replicated with a consistent direction and nominal significance in deCODE, two in the Japan COVID-19 Task Force, and four in All of Us (all *P* < 0.05). The strongest cumulative support was observed for LRRC25, PARK7, and LRRC37A2.

**Conclusions:** These findings prioritize circulating protein candidates for mechanistic, prevention-oriented, and translational follow-up in breast cancer, while underscoring the need for larger ancestrally diverse proteomic resources to support equitable target and biomarker discovery.

## Introduction

Identifying the molecular mediators of breast cancer susceptibility across populations remains a central challenge in cancer biology. Although genome-wide association studies (GWAS) have identified over 230 genetic loci associated with breast cancer risk^1–8^, most risk-associated variants lie in noncoding regions, and their downstream effector mechanisms remain largely unknown^9^. The plasma proteome offers an informative intermediate layer for addressing this gap. Circulating proteins are closer to disease-related biology than germline variants, many are therapeutically tractable, and their levels can be anchored genetically through protein quantitative trait loci (pQTLs)^10–12^. Systematic mapping of genetically predicted protein levels onto breast cancer risk may therefore offer a route from GWAS signals to causal candidates, with implications for risk stratification and therapeutic target identification.

Mendelian randomization (MR) uses genetic variants as instrumental variables (IVs) to estimate the effect of an exposure, such as circulating protein levels, on an outcome^13,14^. Because germline variation is fixed at conception, MR is less susceptible than conventional observational analyses to confounding and reverse causation^15^, and genetically supported targets are more likely to succeed in drug development^14,16^. The potential of proteome-wide MR has expanded substantially with the UK Biobank Pharma Proteomics Project (UKB-PPP), which identified approximately 20 times as many genetic associations with protein levels as prior antibody-based studies^17^. These associations provide instruments of exceptional breadth and strength to support systematic, proteome-scale causal inference.

Previous MR studies of circulating proteins and breast cancer risk have provided valuable insights but faced limitations^18–29^. Some relied on older and smaller pQTL datasets with limited protein coverage. Nearly all were conducted in European-ancestry populations, limiting generalizability and the ability to assess whether genetically predicted protein effects are shared or heterogeneous across ancestries^30,31^. Breast cancer intrinsic-like subtypes differ in molecular characteristics, prognosis, and likely etiology^7,32–34^, yet proteome-wide evidence across subtype contexts remains sparse.

Here, we addressed these gaps through proteome-wide MR analyses of breast cancer risk using *eligible cis*-pQTLs for 1,962 distinct plasma proteins from 34,557 European-ancestry participants in UKB-PPP and breast cancer GWAS data from European-, East Asian-, and African-ancestry participants (156,901 cases and 204,634 controls in total)^6–8,17,35^. We combined ancestry-specific MR estimates through fixed-effects meta-analysis and prioritized 12 candidate proteins associated with overall breast cancer risk. We then evaluated subtype-specific and reverse-direction associations and assessed robustness through a multi-tiered validation framework incorporating statistical colocalization, replication in independent European and East Asian pQTL resources^36,37^, and phenotype-linked genetic validation using protein polygenic scores (PGS) in the ancestrally diverse All of Us cohort (AoU, n = 224,107)^38^. Together, these analyses prioritize plasma proteins linked to breast cancer risk and identify a subset with stronger genetic support for a putative causal role in breast cancer susceptibility.

## Methods

### Overall Study Design

Primary two-sample MR analyses paired European-ancestry UKB-PPP *cis*-pQTL associations for plasma proteins with ancestry-specific breast cancer GWAS associations from European-, East Asian-, and African-ancestry participants. Ancestry-specific MR estimates were subsequently combined via fixed-effects meta-analysis to generate cross-ancestry effect estimates. Fixed-effects meta-analysis was used as the primary discovery framework to test for a non-zero pooled association, while Cochran’s Q statistic was used to test for cross-ancestry heterogeneity. Candidate proteins that reached proteome-wide significance (defined as *P* < 0.05/1,971, where 1,971 is the number of primary tests) in the cross-ancestry meta-analysis and showed robustness across alternative MR estimators and instrument-selection thresholds were carried forward for downstream analyses (**Figure 1**). Candidates were evaluated by combining evidence across validation layers to distinguish proteins with stronger cumulative support from those with more provisional evidence.

**Figure 1.**
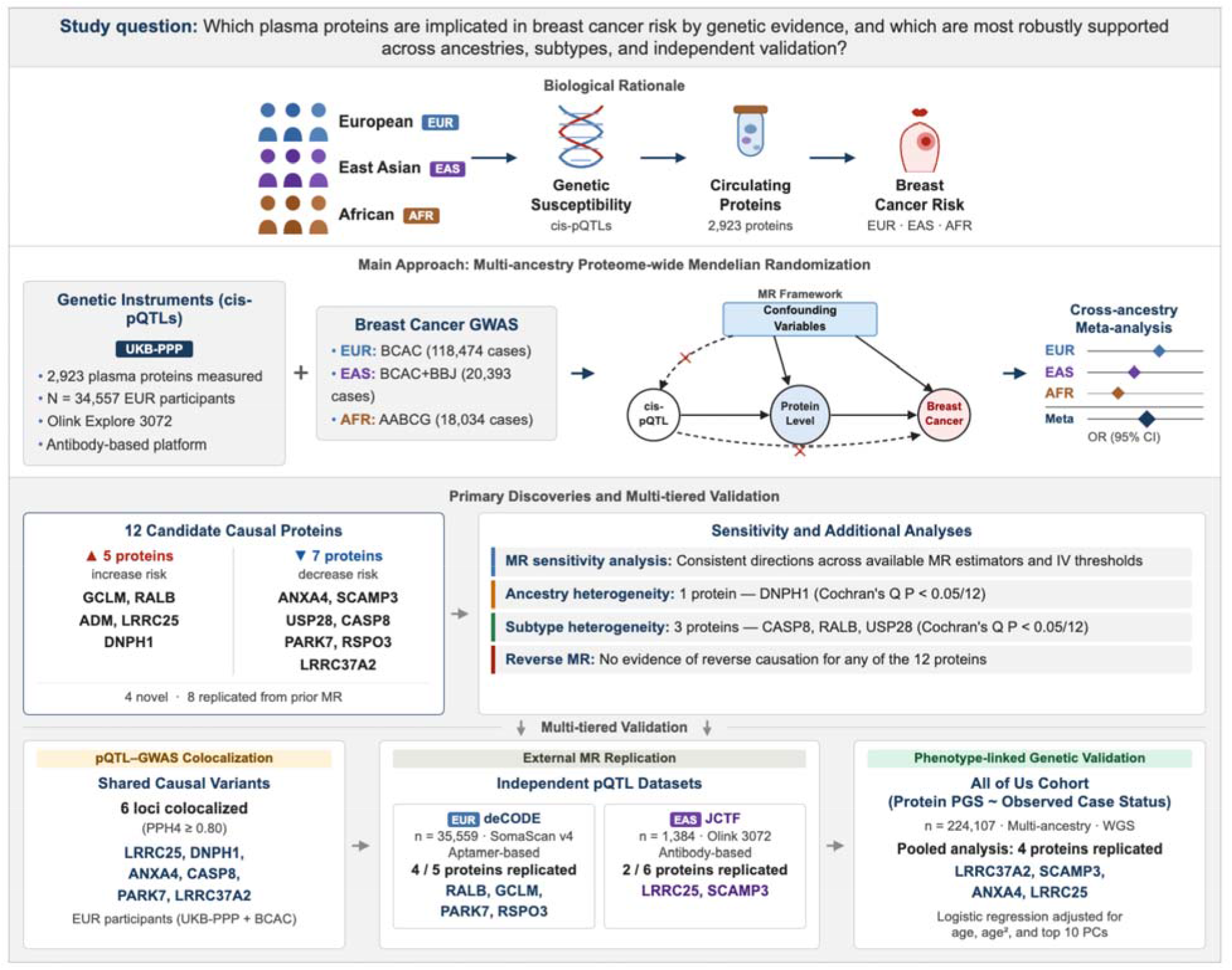
Graphical abstract. We first conducted a proteome-wide Mendelian randomization (MR) study to evaluate 822 associations between genetically predicted plasma protein levels and breast cancer risk. Genetic instruments derived from European-ancestry participants in the UK Biobank Pharma Proteomics Project (UKB-PPP; n = 34,557) were integrated with breast cancer genome-wide association studies (GWAS) from European-, East Asian-, and African ancestry populations (156,901 cases and 204,634 controls). Fixed-effects meta-analysis combining cross-ancestry MR estimates identified 12 candidate proteins associated with breast cancer risk. These candidates were further evaluated using subtype-specific and reverse-direction MR analyses, statistical colocalization, external MR replication in two independent proteomic datasets (deCODE, n = 35,559; Japan COVID-19 Task Force [JCTF], n = 1,384), and phenotype-linked genetic validation in the ancestrally diverse All of Us cohort (n = 224,107). Support varied across candidates, with a subset showing stronger multi-layer evidence. Alt text: Study flowchart linking genetic predictors of plasma protein levels from the UK Biobank with breast cancer data from European, East Asian, and African ancestry groups. Twelve candidate proteins undergo downstream analyses. Support differs across proteins and validation approaches.

### Main Data Sources

#### Protein Data Source

Genetic variant-protein association summary statistics were obtained from the UKB-PPP, which measured 2,923 plasma proteins using the antibody-based Olink Explore 3072 platform^17^ (**Table 1**). Our analysis utilized the discovery pQTL dataset, comprising 34,557 European-ancestry participants drawn from the 46,595 randomly selected baseline participants. The latter group had a mean age of 56.7 years (standard deviation [SD] 8.1), and 54.3% were female. The full-cohort UKB-PPP pQTL dataset (n = 52,363 participants) was also retrieved for complementary analysis. This dataset includes the discovery participants along with additional participants from multiple ancestry groups and non-randomly sampled components^17^.

**Table 1.**
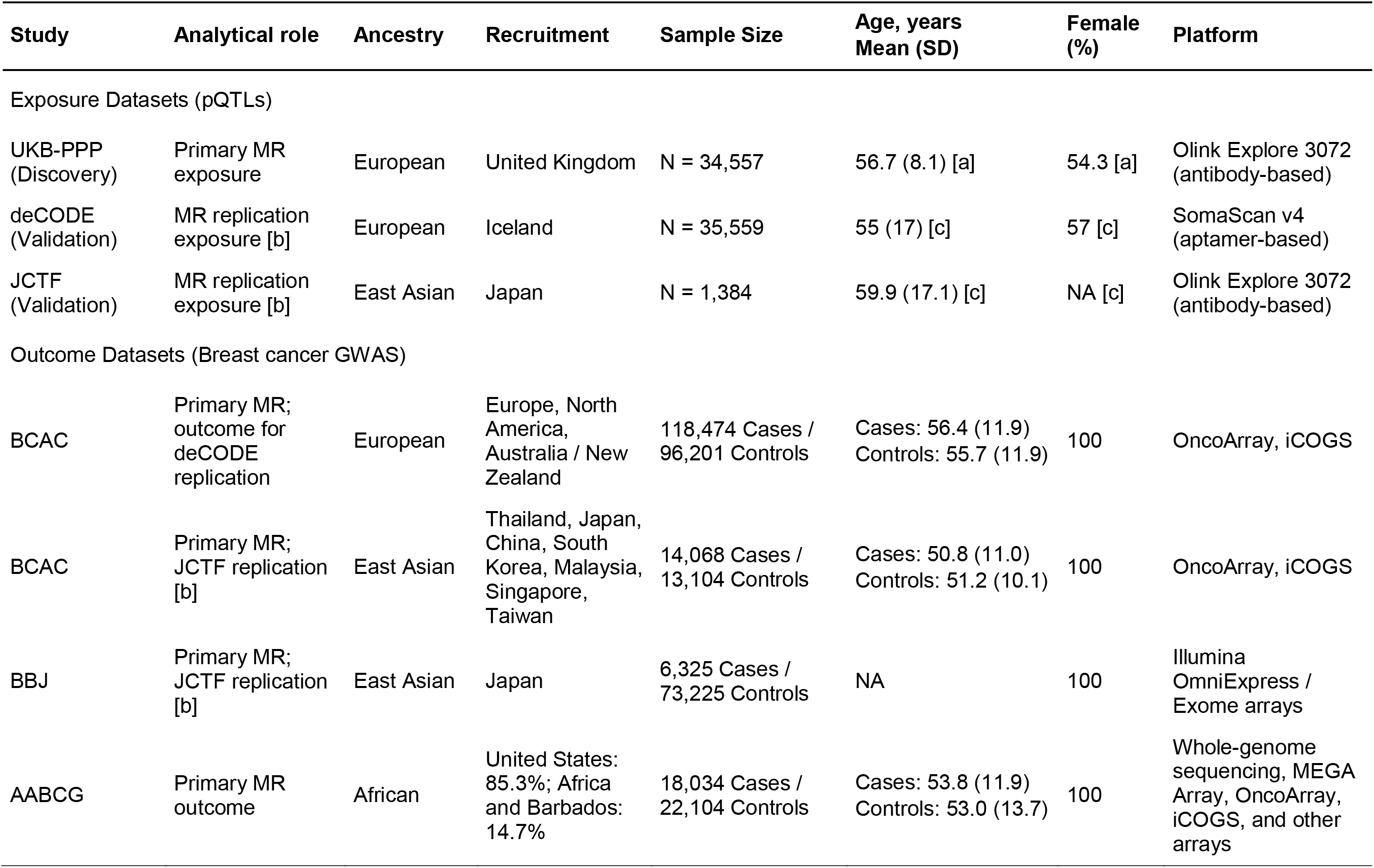
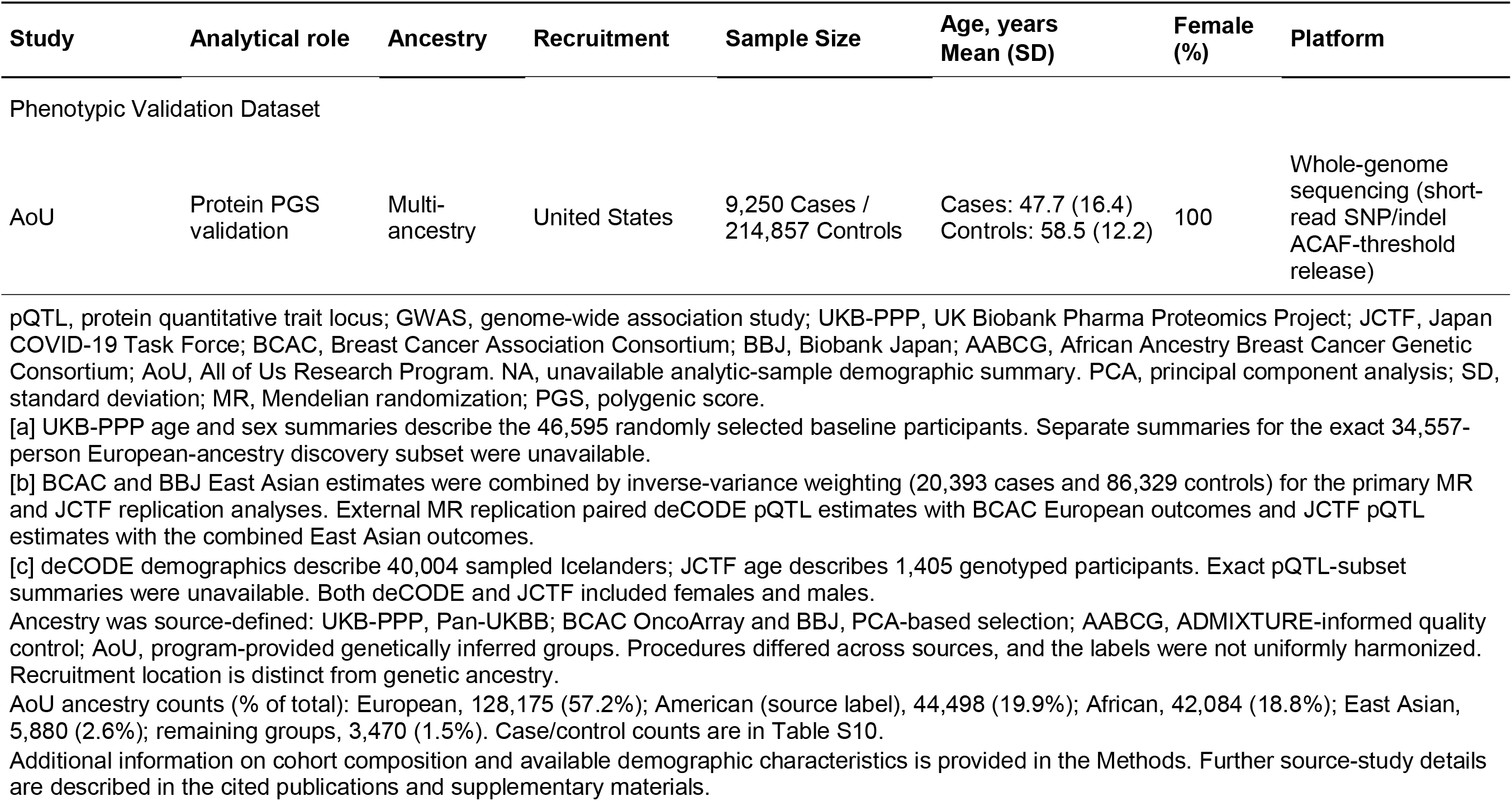
Characteristics of main datasets used in the analyses.

| Study | Analytical role | Ancestry | Recruitment | Sample Size | Age, years<br>Mean (SD) | Female (%) | Platform |
|---|---|---|---|---|---|---|---|
| Exposure Datasets (pQTLs) |  |  |  |  |  |  |  |
| UKB-PPP<br>(Discovery) | Primary MR exposure | European | United Kingdom | N = 34,557 | 56.7 (8.1) [a] | 54.3 [a] | Olink Explore 3072 (antibody-based) |
| deCODE<br>(Validation) | MR replication exposure [b] | European | Iceland | N = 35,559 | 55 (17) [c] | 57 [c] | SomaScan v4 (aptamer-based) |
| JCTF<br>(Validation) | MR replication exposure [b] | East Asian | Japan | N = 1,384 | 59.9 (17.1) [c] | NA [c] | Olink Explore 3072 (antibody-based) |
| Outcome Datasets (Breast cancer GWAS) |  |  |  |  |  |  |  |
| BCAC | Primary MR; outcome for deCODE replication | European | Europe, North America, Australia / New Zealand | 118,474 Cases / 96,201 Controls | Cases: 56.4 (11.9)<br>Controls: 55.7 (11.9) | 100 | OncoArray, iCOGS |
| BCAC | Primary MR; JCTF replication [b] | East Asian | Thailand, Japan, China, South Korea, Malaysia, Singapore, Taiwan | 14,068 Cases / 13,104 Controls | Cases: 50.8 (11.0)<br>Controls: 51.2 (10.1) | 100 | OncoArray, iCOGS |
| BBJ | Primary MR; JCTF replication [b] | East Asian | Japan | 6,325 Cases / 73,225 Controls | NA | 100 | Illumina OmniExpress / Exome arrays |
| AABCG | Primary MR outcome | African | United States: 85.3%; Africa and Barbados: 14.7% | 18,034 Cases / 22,104 Controls | Cases: 53.8 (11.9)<br>Controls: 53.0 (13.7) | 100 | Whole-genome sequencing, MEGA Array, OncoArray, iCOGS, and other arrays |
| Study | Analytical role | Ancestry | Recruitment | Sample Size | Age, years<br>Mean (SD) | Female (%) | Platform |
| Phenotypic Validation Dataset |  |  |  |  |  |  |  |
| AoU | Protein PGS validation | Multi-ancestry | United States | 9,250 Cases / 214,857 Controls | Cases: 47.7 (16.4)<br>Controls: 58.5 (12.2) | 100 | Whole-genome sequencing (short-read SNP/indel ACAF-threshold release) |

pQTL, protein quantitative trait locus; GWAS, genome-wide association study; UKB-PPP, UK Biobank Pharma Proteomics Project; JCTF, Japan COVID-19 Task Force; BCAC, Breast Cancer Association Consortium; BBJ, Biobank Japan; AABCG, African Ancestry Breast Cancer Genetic Consortium; AoU, All of Us Research Program. NA, unavailable analytic-sample demographic summary. PCA, principal component analysis; SD, standard deviation; MR, Mendelian randomization; PGS, polygenic score.
[a] UKB-PPP age and sex summaries describe the 46,595 randomly selected baseline participants. Separate summaries for the exact 34,557-person European-ancestry discovery subset were unavailable.
[b] BCAC and BBJ East Asian estimates were combined by inverse-variance weighting (20,393 cases and 86,329 controls) for the primary MR and JCTF replication analyses. External MR replication paired deCODE pQTL estimates with BCAC European outcomes and JCTF pQTL estimates with the combined East Asian outcomes.
[c] deCODE demographics describe 40,004 sampled Icelanders; JCTF age describes 1,405 genotyped participants. Exact pQTL-subset summaries were unavailable. Both deCODE and JCTF included females and males.
Ancestry was source-defined: UKB-PPP, Pan-UKBB; BCAC OncoArray and BBJ, PCA-based selection; AABCG, ADMIXTURE-informed quality control; AoU, program-provided genetically inferred groups. Procedures differed across sources, and the labels were not uniformly harmonized. Recruitment location is distinct from genetic ancestry.
AoU ancestry counts (% of total): European, 128,175 (57.2%); American (source label), 44,498 (19.9%); African, 42,084 (18.8%); East Asian, 5,880 (2.6%); remaining groups, 3,470 (1.5%). Case/control counts are in Table S10.
Additional information on cohort composition and available demographic characteristics is provided in the Methods. Further source-study details are described in the cited publications and supplementary materials.

Blood samples for the discovery cohort were collected at the UKB baseline recruitment visit (2006–2010) and were subsequently assayed in 2021–2022. Protein levels were measured as normalized protein expression (NPX) values on a log2 scale, then inverse-rank normalized and adjusted for age, age squared, sex, age-by-sex terms, batch, UKB assessment center, UKB genotyping array, time between blood sampling and protein measurement, and the first 20 genetic principal components, before genetic variant–protein association testing. Genetic ancestry was defined by the UKB-PPP research group using pan-UKBB ancestry definitions, in which participants were projected into a principal-component space derived from 1000 Genomes Project (1KGP) and Human Genome Diversity Panel reference samples and classified into broad continental genetic-ancestry groups. Although smaller African-(n = 931) and East Asian-ancestry (n = 262) pQTL datasets were available within UKB-PPP, their sample sizes were insufficient for proteome-wide instrument discovery.

#### Breast Cancer GWAS Data Sources

Breast cancer GWAS summary statistics were obtained from large, ancestry-stratified consortia, all restricted to females (**Table 1**).

European-ancestry summary statistics comprised 118,474 cases (mean age 56.4 years, SD 11.9) and 96,201 controls (mean age 55.7 years, SD 11.9) from 82 studies in the Breast Cancer Association Consortium (BCAC) combining iCOGS and OncoArray, with recruitment predominantly in Europe, North America, and Australia/New Zealand^7^. East Asian-ancestry summary statistics combined BCAC (14,068 cases, mean age 50.8 years, SD 11.0; 13,104 controls, mean age 51.2 years, SD 10.1), with contributing studies in Thailand, Japan, China, South Korea, Malaysia, Singapore, and Taiwan, and BioBank Japan (BBJ; 6,325 cases; 73,225 controls; age summaries unavailable), which recruited predominantly Japanese participants through medical institutions throughout Japan^6,35,39^. African-ancestry summary statistics comprised 18,034 cases (mean age 53.8 years, SD 11.9) and 22,104 controls (mean age 53.0 years, SD 13.7) from 26 studies in the African Ancestry Breast Cancer Genetic Consortium (AABCG); 85.3% of participants were recruited in the United States and 14.7% in Africa and Barbados^8^.

Ancestry labels for the breast cancer outcome GWAS were adopted directly from the source BCAC, BBJ, and AABCG studies, with ancestry classification and quality control performed internally by each consortium prior to release. Detailed study composition and ancestry criteria are reported in the original publications.

European-ancestry summary statistics for five intrinsic-like breast cancer subtypes were additionally drawn from BCAC: luminal A-like, luminal B/HER2-negative-like, luminal B-like, HER2-enriched-like, and triple-negative breast cancer (TNBC)^7^.

These analyses included 27,481, 6,799, 6,502, 2,793, and 7,174 observed cases, respectively, with 91,477 shared controls. We accounted for the shared controls when comparing results across subtypes, as described in **Supplementary Methods**.

### Ethics approval and consent to participate

This study used published summary-level genome-wide association study data and deidentified individual-level data from the All of Us Research Program. Ethical approvals and participant consent procedures for the contributing studies are described in their original publications. The All of Us Research Program was approved by its Institutional Review Board, and participants provided informed consent for research use of their data. All of Us analyses were conducted within the Researcher Workbench using Controlled Tier data. The All of Us Institutional Review Board determined that research using Controlled Tier data does not constitute human subjects research and does not require review of individual projects by that board (determination 2021-02-TN-001).

## Statistical Analysis

### Mendelian Randomization Analysis

#### MR assumptions

MR estimates were interpreted under three core IV conditions. Relevance requires that the selected *cis*-pQTL instruments are strongly associated with plasma protein levels. Independence requires that the association between the instruments and breast cancer is not confounded, for example, by population stratification or other unmeasured factors. Exclusion restriction requires that the instruments influence breast cancer only through the target protein, and not through other, independent pathways (horizontal pleiotropy). We evaluated the plausibility of these assumptions via strict instrument-selection criteria, alternative MR estimators that relax different assumptions about pleiotropy, and additional complementary analyses including colocalization. These analyses, however, cannot establish instrument validity or exclude residual pleiotropy. Throughout the forward MR analyses, odds ratios (ORs) represent breast cancer odds per 1-SD increase in genetically predicted plasma protein levels. We use “risk” to refer to breast cancer susceptibility.

#### Data harmonization and instrumental variable selection

European-ancestry pQTL summary statistics were harmonized against each ancestry-specific breast cancer GWAS. Only variants present in both datasets were retained. Effect allele alignment was verified between datasets, and effect sizes were multiplied by −1 where necessary to align directions across variants. Within each harmonized variant set, *cis*-acting pQTLs were defined as variants within ±1 Mb of the protein-coding gene, excluding variants on sex chromosomes or those with imputation quality below threshold (INFO score < 0.3) or low minor allele frequency (MAF < 0.01). Independent *cis*-pQTLs were then identified via linkage disequilibrium (LD) clumping (r² ≤ 0.01 within a 2 Mb window) using the 1KGP European-ancestry reference panel^40^, retaining variants at a genome-wide significant threshold (*P* < 5 × 10^-8^) for primary analyses; relaxed thresholds used in sensitivity analyses are described below. Because harmonization was performed independently for each ancestry-specific GWAS before LD clumping, the final IV sets differ slightly across ancestry-specific analyses, reflecting variation in variant availability across GWAS datasets. Proteins yielding no eligible instruments at *P* < 5 × 10^-8^ after harmonization in a given ancestry were excluded from that analysis. The strength of each selected *cis-pQTL* as an instrument for plasma protein level was assessed using the approximate *F*-statistic, calculated as the square of the ratio of the UKB-PPP variant-protein effect estimate to its standard error.

#### Primary and sensitivity MR analyses

Primary MR analyses proceeded in two steps. First, ancestry-specific MR was conducted using the inverse-variance weighted (IVW) method^13^ with instruments selected at *P* < 5 × 10^-8^. Second, estimates available from two or more ancestry-specific analyses were combined via fixed-effects meta-analysis; when only one ancestry-specific estimate was available, it was retained in the primary analysis. Statistical significance was defined by a Bonferroni-corrected threshold based on the number of primary tests (*P* < 0.05/1,971). Proteins measured on multiple panels were analyzed separately for each panel, with each protein–panel combination counted as a separate test. Cross-ancestry heterogeneity was assessed using Cochran’s Q test, with the significance threshold Bonferroni-corrected for the number of significant candidate proteins (*P* < 0.05/12). IVW MR analyses used the mr_ivw function in the MendelianRandomization package with first-order (“simple”) weights and a fixed-effects model for two or three instruments or a multiplicative random-effects model for four or more instruments. Single-instrument analyses used the Wald ratio with a first-order standard error.

Sensitivity MR analyses were conducted by replacing IVW with alternative MR methods in the ancestry-specific analyses to evaluate robustness to horizontal pleiotropy and weak instrument bias, including the weighted median method^41^, which provides consistent estimates when more than 50% of the total weight is contributed by valid instruments; the penalized IVW method (pIVW)^42^, which corrects for weak instrument bias; and MR-Robust Adjusted Profile Score (MR-RAPS)^43^, which accounts for both systematic and idiosyncratic pleiotropy. Weighted median analyses used the same genome-wide significant instruments as the IVW analysis. pIVW and MR-RAPS analyses were additionally evaluated using relaxed IV selection thresholds (*P* < 5 × 10^-7^ and *P* < 5 × 10^-6^) to assess robustness to instrument selection criteria. A protein was considered a significant candidate if its MR association reached Bonferroni-corrected significance in the primary fixed-effects meta-analysis and showed a consistent direction across all available fixed-effects meta-analyses using alternative MR methods.

To assess sensitivity to the exposure dataset, we repeated the primary cross-ancestry MR pipeline using the full-cohort UKB-PPP pQTL dataset (n = 52,363), reselecting IVs under the original criteria. We retained the European-ancestry LD reference because European-ancestry participants predominated in this cohort.

We conducted three additional sensitivity analyses for the significant prioritized proteins. First, we evaluated cross-ancestry associations using random-effects meta-analysis with restricted maximum likelihood (REML) estimation and the Han and Eskin RE2 test implemented in Metasoft^44^ (**Supplementary Methods**). Second, we assessed sensitivity to sex-combined pQTL associations using female-only pQTL estimates from 25,904 European-ancestry UKB-PPP females^45^. For each protein, we retained the instruments from the primary European-ancestry MR analysis that were available in the female-only dataset. Using this identical instrument subset and the same European-ancestry BCAC outcome associations, we compared MR estimates calculated with female-only versus sex-combined pQTL associations. Third, we assessed sensitivity to the East Asian outcome source by repeating MR using BCAC East Asian and BBJ summary statistics separately, retaining UKB-PPP discovery pQTLs as the exposure dataset.

For DNPH1, which demonstrated statistically significant cross-ancestry heterogeneity, a targeted sensitivity analysis was performed to assess whether the heterogeneity was attributable to European-ancestry-derived instrument weights. For the genome-wide significant *cis*-pQTLs used as instruments in the primary East Asian-specific MR, we extracted their effect estimates from the East Asian-ancestry UKB-PPP pQTL dataset (n = 262) and used them as weights to re-run IVW MR against the East Asian-ancestry breast cancer GWAS.

#### Subtype-specific and reverse MR analyses

For the significant candidate proteins, additional IVW MR analyses restricted to European-ancestry participants were performed. For subtype-specific MR, the same genome-wide significant *cis*-pQTL instruments and variant-protein effect estimates were used, with instrument-outcome associations extracted from the summary statistics of each subtype-specific breast cancer GWAS. Heterogeneity across subtypes was evaluated using a generalized Cochran’s Q test with a covariance matrix accounting for shared controls (**Supplementary Methods**). For reverse MR, independent genome-wide significant variants (*P* < 5 × 10^-8^) for overall breast cancer were selected from the European-ancestry BCAC GWAS after harmonization with the UKB-PPP discovery pQTL data, using the same variant filtering and LD-clumping criteria as in the forward analyses. For both analyses, statistical significance was determined using a Bonferroni-corrected threshold based on the number of proteins tested.

Normal-based association tests were two-sided; Cochran’s Q heterogeneity tests used upper-tail chi-square probabilities. MR analyses were conducted in R using the MendelianRandomization, mr.raps, mr.pIVW, and meta packages.

### Multi-layer Validation

#### Colocalization analysis

To strengthen causal inference, colocalization analyses were performed to test whether the observed MR associations were driven by a shared causal variant influencing both plasma protein levels and breast cancer risk. For each candidate protein, analysis regions were defined as ±150 kb windows around each lead *cis*-pQTL instrument. For proteins with multiple instruments, colocalization was performed independently within each instrument’s analysis region. Within each region, posterior probabilities for five mutually exclusive hypotheses (H0–H4) were calculated using the coloc R package with default prior probabilities^46^. Evidence for colocalization was evaluated based on the posterior probability of hypothesis 4 (PPH4), which quantifies support for a single shared causal variant. A protein was considered to show strong evidence of colocalization if PPH4 ≥ 0.80 in any tested region, and the highest PPH4 across regions is reported. For regions meeting this threshold, SNP.PP.H4 denotes the posterior probability that a particular variant is the shared causal variant, conditional on H4 being true. To use ancestry-matched exposure and outcome data, analyses were restricted to European-ancestry participants, using the European-ancestry UKB-PPP discovery pQTL dataset for protein levels and BCAC summary statistics for overall breast cancer risk.

#### External MR replication using independent pQTL resources

External MR replication was performed for each candidate protein using independent pQTL datasets from deCODE (n = 35,559 Icelandic participants; SomaScan v4 aptamer-based platform)^36^ and the Japan COVID-19 Task Force (JCTF; n = 1,384 Japanese participants; Olink Explore 3072 antibody-based platform)^37^ (**Table 1**). Published demographic summaries describe the broader source samples: deCODE reported a mean age of 55 years (SD 17) and 57% female among 40,004 sampled Icelanders, and JCTF reported a mean age of 59.9 years (SD 17.1) among 1,405 genotyped participants. For each dataset, independent genetic instruments were re-identified using the same LD clumping procedure applied in the primary analysis, with ancestry-matched 1KGP reference panels used for LD estimation. A genome-wide significance threshold of *P* < 5 × 10^-8^ was applied for instrument selection in deCODE, while a relaxed threshold of *P* < 5 × 10^-7^ was used for JCTF to accommodate its more limited sample size. Instrument-outcome association estimates were extracted from the primary European-and East Asian-ancestry overall breast cancer GWAS for deCODE and JCTF analyses, respectively, and IVW MR was performed using these instruments and their ancestry-specific effect estimates. Supportive replication was defined as a directionally consistent association achieving nominal significance (*P* < 0.05).

#### Phenotype-linked genetic validation in the All of Us cohort

To evaluate whether inherited genetic predisposition to altered levels of each candidate protein is associated with breast cancer risk in an independent, ancestrally diverse cohort, we constructed per-protein PGS and tested their associations with observed breast cancer status in females from AoU Controlled Tier v8^38^ (**Table 1**).

Genetic data were derived from the whole-genome sequencing short-read SNP/indel ACAF-threshold release. Each PGS was computed as the allele-weighted sum of *cis*-pQTLs used in the corresponding European-ancestry primary MR analysis, with weights defined by their *cis*-pQTL effect estimates from the UKB-PPP discovery dataset. Breast cancer case status was defined using a curated set of Observational Medical Outcomes Partnership (OMOP) condition_concept_id entries representing breast cancer-related diagnoses derived from breast-related malignant neoplasm concepts. Inferred genetic ancestry labels were used as provided by the AoU Research Program. Full details of the ancestry inference procedure are described in the AoU genomic data quality report. For cases, age at breast cancer diagnosis was assigned as the earliest age across qualifying condition records; controls retained their baseline age. The mean age at diagnosis was 47.7 years (SD 16.4) among cases, and the mean baseline age was 58.5 years (SD 12.2) among controls (**Table 1**).

Analyses were conducted in the multi-ancestry pooled sample and subsequently stratified by inferred genetic ancestry. Before each analysis, PGSs were standardized to unit variance within the corresponding analytic sample. The association between each standardized PGS and breast cancer status was evaluated using logistic regression, adjusting for age, age squared, and the top 10 genetic principal components. Nominal significance (*P* < 0.05) was used as the replication threshold.

## Results

### Proteome-wide cross-ancestry meta-analysis

**Table 1** summarizes the sample sizes, available demographic characteristics, recruitment locations, and analytical roles of the main datasets. After data harmonization and exclusion of proteins yielding no eligible *cis*-pQTLs, the primary analysis included 1,971 tests representing 1,962 distinct proteins. Of these tests, 1,955 had MR estimates from at least two ancestry groups, which were combined by fixed-effects meta-analysis; 16 had only European-ancestry estimates, which were retained in the primary analysis. All selected *cis*-pQTLs had *F*-statistics > 10, suggesting a low risk of weak instrument bias (**Table S1).**

The primary IVW analysis identified 12 plasma proteins (0.61% of the 1,971 tests) associated with overall breast cancer risk, with *P*-values below the Bonferroni threshold of 0.05/1,971 (**Figure 2**; **Table 2**). Higher genetically predicted levels of five proteins (GCLM, RALB, ADM, LRRC25, DNPH1) were associated with higher odds of breast cancer, while higher genetically predicted levels of seven proteins (ANXA4, SCAMP3, USP28, CASP8, PARK7, RSPO3, LRRC37A2) were associated with lower odds. Of these, eight proteins corroborated significant MR findings defined and reported by previous studies^23,25–29^, whereas four (GCLM, LRRC25, ANXA4, USP28) had not, to our knowledge, been previously prioritized by MR studies of breast cancer risk. All 12 associations were directionally consistent across available sensitivity analyses using alternative MR estimators and relaxed instrument-selection thresholds (**Figure S1**, **Table S2**). These 12 candidate proteins were carried forward for downstream analyses.

**Figure 2.**
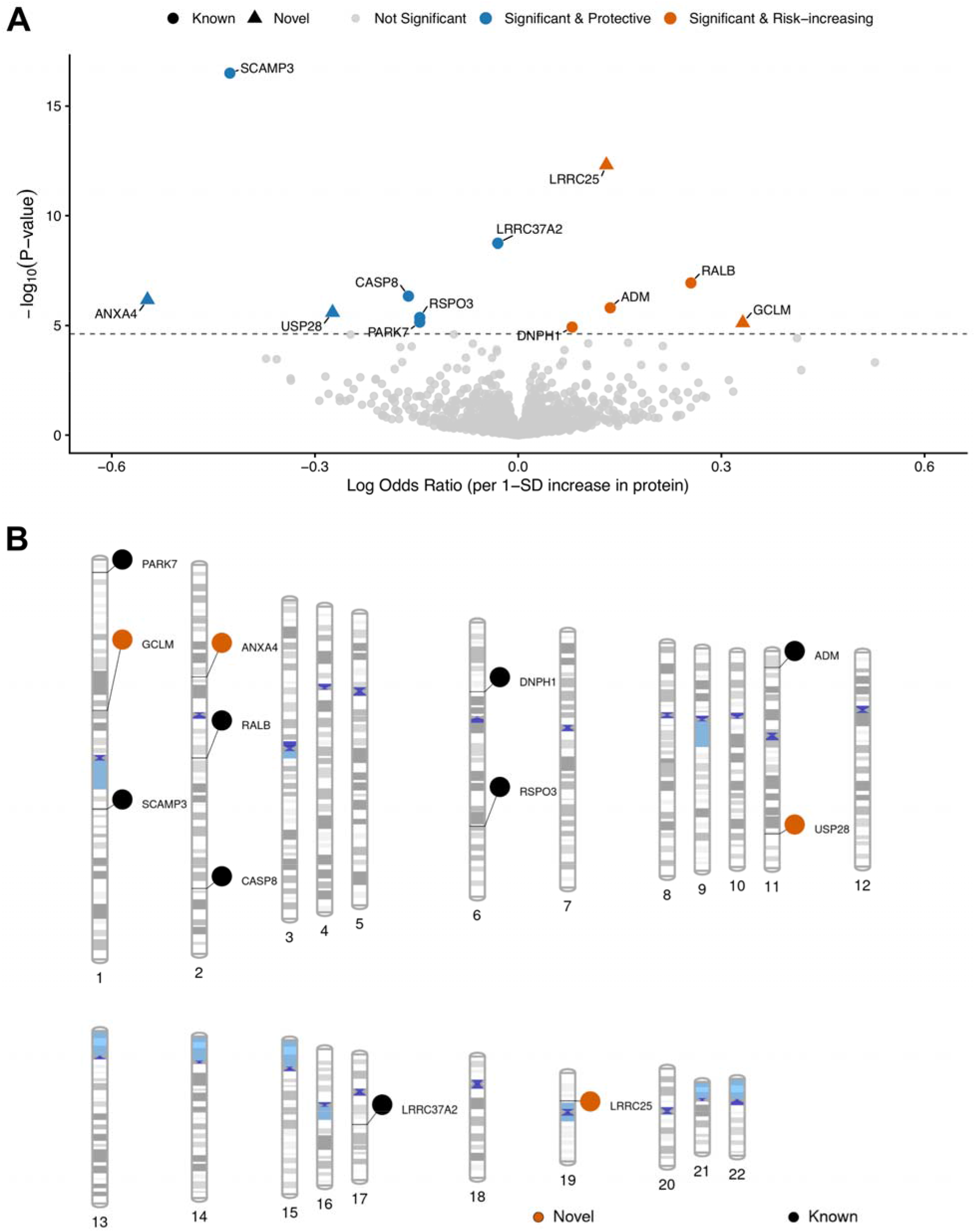
Volcano plot and genomic mapping of 12 candidate plasma proteins associated with overall breast cancer risk. (A) Volcano plot of cross-ancestry meta-analysis estimates for genetically predicted plasma protein levels in relation to overall breast cancer risk. The x-axis shows the log-transformed odds ratio (log OR) per 1-standard deviation increase in genetically predicted protein levels, and the y-axis shows statistical significance as the negative log_10_-transformed *P*-value. The horizontal dashed line indicates the Bonferroni-corrected significance threshold (*P* < 0.05/1,971). Of the 12 significant proteins, eight corroborated associations reported by prior MR studies (circles), and four were not previously prioritized by MR (triangles). Novelty is defined relative to prior MR literature and does not imply these proteins have not been studied in breast cancer biology through other approaches. (B) Phenogram depicting the chromosomal locations of genes encoding the 12 candidate proteins. Alt text: Panel A plots genetic association estimates for plasma proteins against statistical significance. Twelve proteins meet the discovery threshold, with five associated with higher breast cancer odds and seven with lower odds. Panel B marks the chromosomal locations of the corresponding genes. Symbols distinguish previously reported candidates from newly prioritized candidates.

**Table 2.**
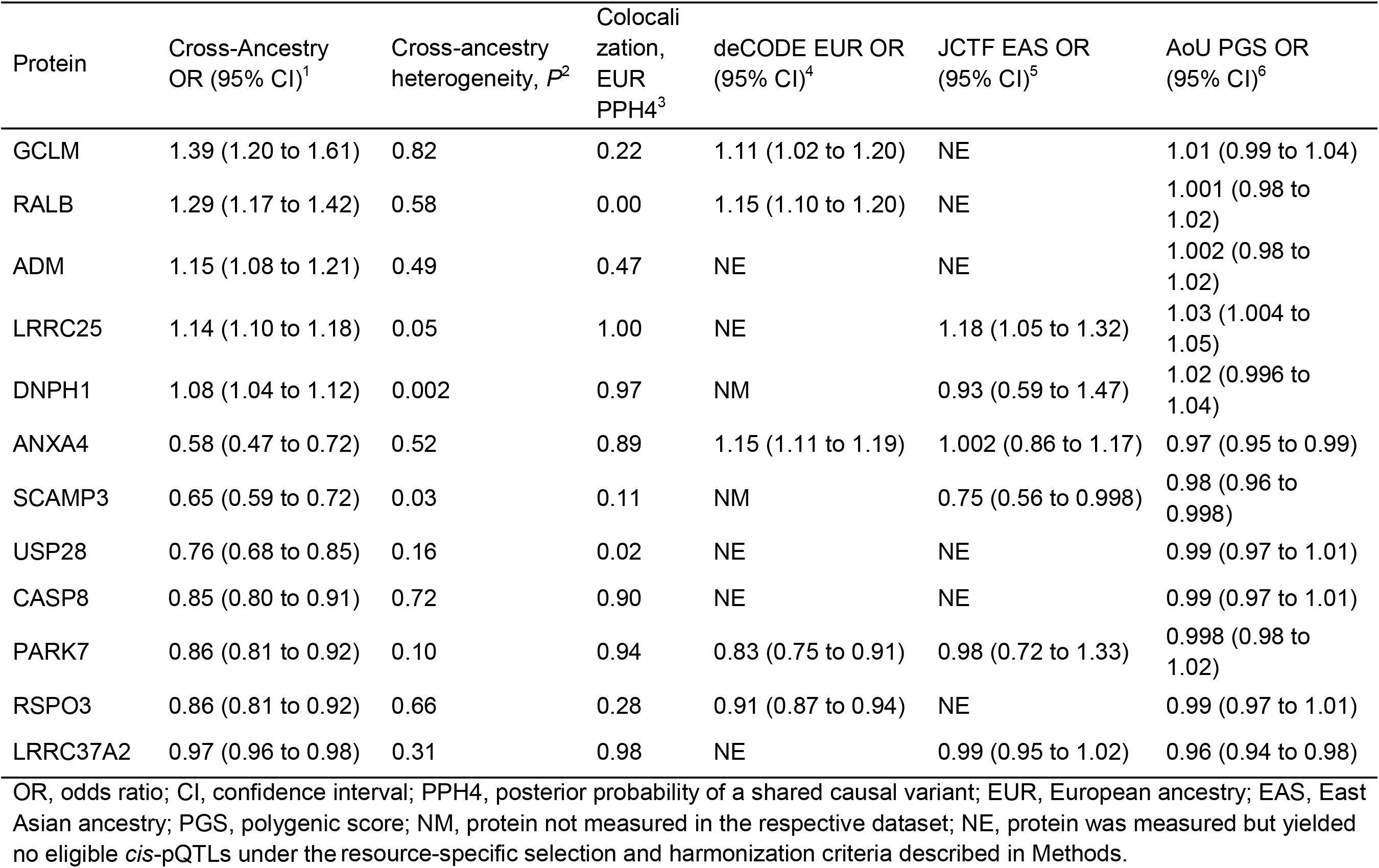
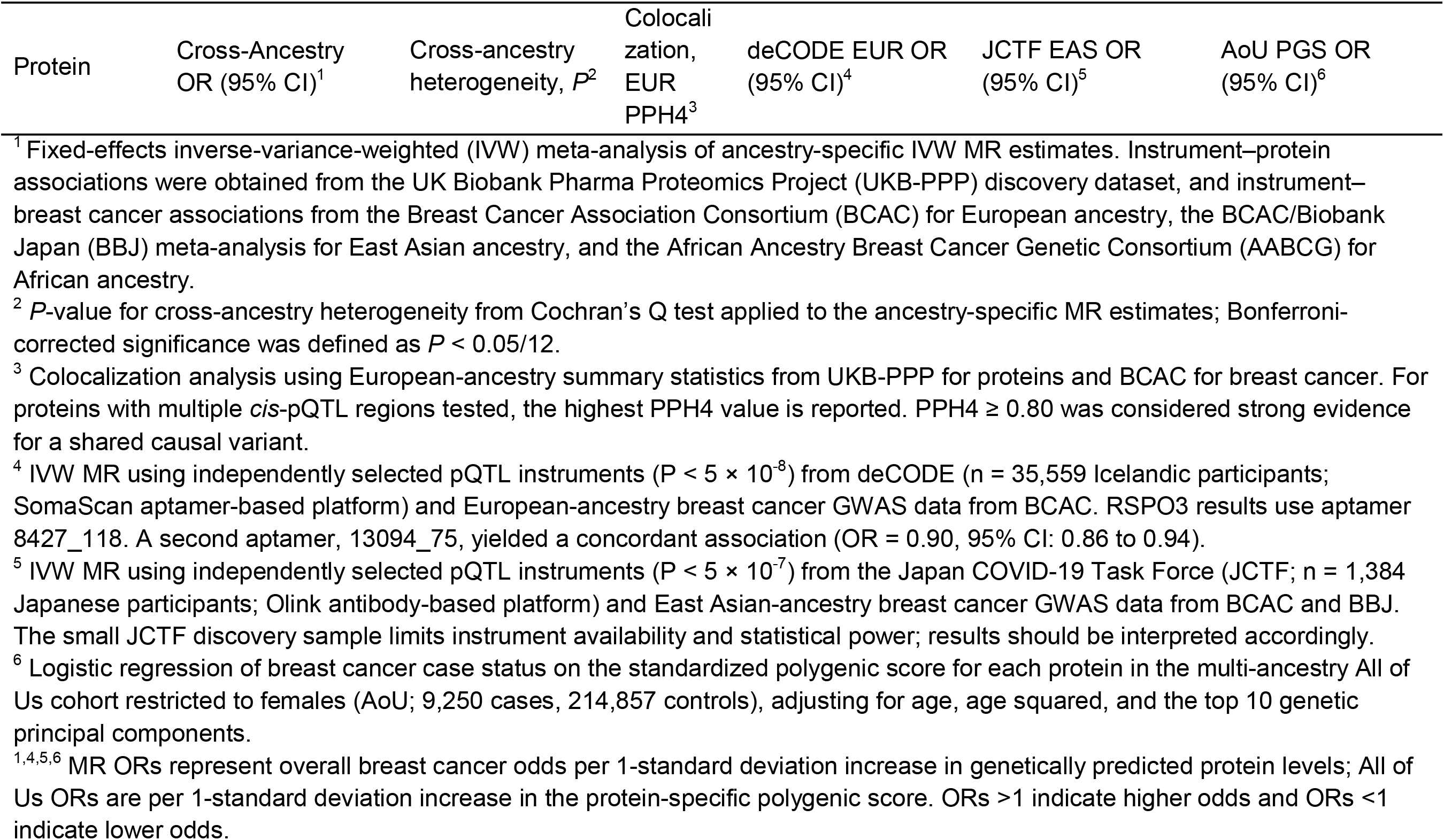
Summary of multi-tiered validation evidence for 12 candidate plasma proteins associated with breast cancer risk.

| Protein | Cross-Ancestry OR (95% CI)<sup>1</sup> | Cross-ancestry heterogeneity, *P*<sup>2</sup> | Colocalization, EUR PPH4<sup>3</sup> | deCODE EUR OR (95% CI)<sup>4</sup> | JCTF EAS OR (95% CI)<sup>5</sup> | AoU PGS OR (95% CI)<sup>6</sup> |
|---|---|---|---|---|---|---|
| GCLM | 1.39 (1.20 to 1.61) | 0.82 | 0.22 | 1.11 (1.02 to 1.20) | NE | 1.01 (0.99 to 1.04) |
| RALB | 1.29 (1.17 to 1.42) | 0.58 | 0.00 | 1.15 (1.10 to 1.20) | NE | 1.001 (0.98 to 1.02) |
| ADM | 1.15 (1.08 to 1.21) | 0.49 | 0.47 | NE | NE | 1.002 (0.98 to 1.02) |
| LRRC25 | 1.14 (1.10 to 1.18) | 0.05 | 1.00 | NE | 1.18 (1.05 to 1.32) | 1.03 (1.004 to 1.05) |
| DNPH1 | 1.08 (1.04 to 1.12) | 0.002 | 0.97 | NM | 0.93 (0.59 to 1.47) | 1.02 (0.996 to 1.04) |
| ANXA4 | 0.58 (0.47 to 0.72) | 0.52 | 0.89 | 1.15 (1.11 to 1.19) | 1.002 (0.86 to 1.17) | 0.97 (0.95 to 0.99) |
| SCAMP3 | 0.65 (0.59 to 0.72) | 0.03 | 0.11 | NM | 0.75 (0.56 to 0.998) | 0.98 (0.96 to 0.998) |
| USP28 | 0.76 (0.68 to 0.85) | 0.16 | 0.02 | NE | NE | 0.99 (0.97 to 1.01) |
| CASP8 | 0.85 (0.80 to 0.91) | 0.72 | 0.90 | NE | NE | 0.99 (0.97 to 1.01) |
| PARK7 | 0.86 (0.81 to 0.92) | 0.10 | 0.94 | 0.83 (0.75 to 0.91) | 0.98 (0.72 to 1.33) | 0.998 (0.98 to 1.02) |
| RSPO3 | 0.86 (0.81 to 0.92) | 0.66 | 0.28 | 0.91 (0.87 to 0.94) | NE | 0.99 (0.97 to 1.01) |
| LRRC37A2 | 0.97 (0.96 to 0.98) | 0.31 | 0.98 | NE | 0.99 (0.95 to 1.02) | 0.96 (0.94 to 0.98) |

OR, odds ratio; CI, confidence interval; PPH4, posterior probability of a shared causal variant; EUR, European ancestry; EAS, East Asian ancestry; PGS, polygenic score; NM, protein not measured in the respective dataset; NE, protein was measured but yielded no eligible *cis*-pQTLs under the resource-specific selection and harmonization criteria described in Methods.
<sup>1</sup> Fixed-effects inverse-variance-weighted (IVW) meta-analysis of ancestry-specific IVW MR estimates. Instrument–protein associations were obtained from the UK Biobank Pharma Proteomics Project (UKB-PPP) discovery dataset, and instrument–breast cancer associations from the Breast Cancer Association Consortium (BCAC) for European ancestry, the BCAC/Biobank Japan (BBJ) meta-analysis for East Asian ancestry, and the African Ancestry Breast Cancer Genetic Consortium (AABCG) for African ancestry.
<sup>2</sup> *P*-value for cross-ancestry heterogeneity from Cochran's Q test applied to the ancestry-specific MR estimates; Bonferroni-corrected significance was defined as *P* < 0.05/12.
<sup>3</sup> Colocalization analysis using European-ancestry summary statistics from UKB-PPP for proteins and BCAC for breast cancer. For proteins with multiple *cis*-pQTL regions tested, the highest PPH4 value is reported. PPH4 ≥ 0.80 was considered strong evidence for a shared causal variant.
<sup>4</sup> IVW MR using independently selected pQTL instruments (P < 5 × 10<sup>-8</sup>) from deCODE (n = 35,559 Icelandic participants; SomaScan aptamer-based platform) and European-ancestry breast cancer GWAS data from BCAC. RSPO3 results use aptamer 8427\_118. A second aptamer, 13094\_75, yielded a concordant association (OR = 0.90, 95% CI: 0.86 to 0.94).
<sup>5</sup> IVW MR using independently selected pQTL instruments (P < 5 × 10<sup>-7</sup>) from the Japan COVID-19 Task Force (JCTF; n = 1,384 Japanese participants; Olink antibody-based platform) and East Asian-ancestry breast cancer GWAS data from BCAC and BBJ. The small JCTF discovery sample limits instrument availability and statistical power; results should be interpreted accordingly.
<sup>6</sup> Logistic regression of breast cancer case status on the standardized polygenic score for each protein in the multi-ancestry All of Us cohort restricted to females (AoU; 9,250 cases, 214,857 controls), adjusting for age, age squared, and the top 10 genetic principal components.
<sup>1,4,5,6</sup> MR ORs represent overall breast cancer odds per 1-standard deviation increase in genetically predicted protein levels; All of Us ORs are per 1-standard deviation increase in the protein-specific polygenic score. ORs >1 indicate higher odds and ORs <1 indicate lower odds.

Using the full-cohort UKB-PPP pQTL dataset, all 12 primary candidates retained consistent effect directions, and eight met the Bonferroni threshold of *P* < 0.05/2,040 (full results across 2,040 tests are shown in **Table S3**). Seven additional proteins (BRAP, ERP29, EGLN1, RALY, PDIA3, PKD2, and TLR1) met the Bonferroni threshold and were considered exploratory given the sampling variability and cohort composition.

Cross-ancestry sensitivity analyses showed that six of the 12 candidates retained significance under REML-based random-effects meta-analysis, whereas all 12 remained significant under the Han and Eskin RE2 test (*P* < 0.05/1,971; **Table S4).**

Female-only analyses included 66 of the 84 original instruments, covering 11 candidates; RALB could not be evaluated. All 11 estimates were directionally consistent with and highly correlated with the matched sex-combined estimates (Pearson r = 0.9903), and ten had *P* < 0.05/12 (**Table S5**). RSPO3 had the same OR in both analyses (0.871) but did not meet this significance threshold in the female-only analysis.

### Ancestry-specific associations and cross-ancestry heterogeneity

Among the 12 candidate proteins, 10 (all except RSPO3 and GCLM) achieved Bonferroni-corrected significance (*P* < 0.05/1,971) in the European-ancestry MR analysis (**Figure 3**; **Table S6).** None reached Bonferroni-corrected significance in the East Asian-or African-specific analyses, although LRRC25, DNPH1, and ANXA4 were nominally significant (*P* < 0.05) in the East Asian analysis. This pattern probably reflects the substantially larger European-ancestry breast cancer GWAS, together with potentially reduced transferability of European-derived pQTL instruments to non-European participants.

**Figure 3.**
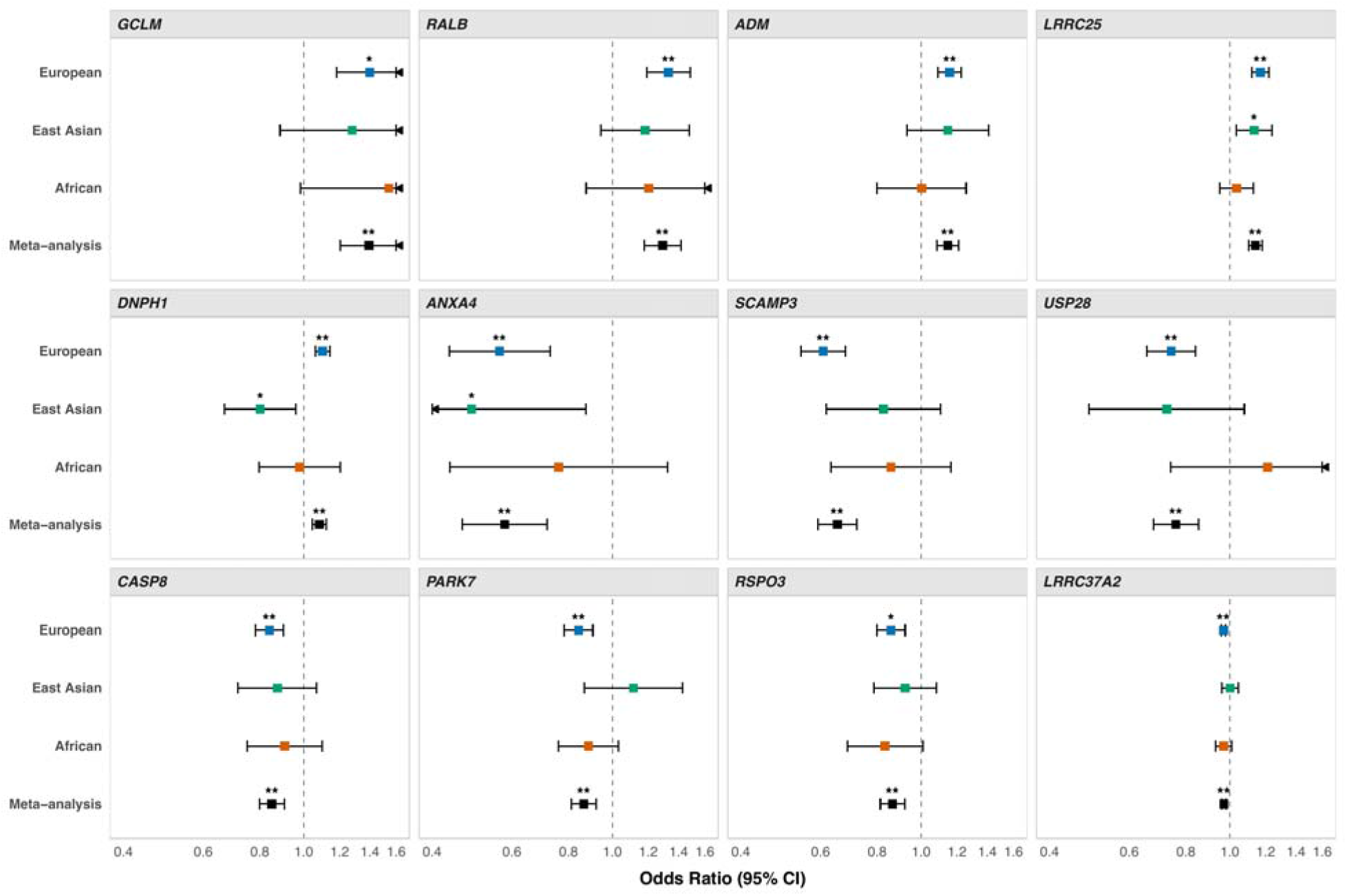
Forest plots of ancestry-specific and cross-ancestry MR estimates for 12 candidate plasma proteins in relation to overall breast cancer risk. Each point represents the odds ratio (OR) per 1-standard deviation increase in genetically predicted protein levels, and horizontal lines indicate 95% confidence intervals (CIs). Results are shown for the European, East Asian, and African ancestry-specific analyses and for the overall fixed-effects cross-ancestry meta-analysis. The x-axis is displayed on a linear scale and truncated at OR = 0.4 and OR = 1.6 for visual clarity; arrows indicate CIs extending beyond this range. Double asterisks (**) denote P < 0.05/1,971, and single asterisks (*) denote nominal significance (0.05/1,971 < *P* < 0.05). Exact effect estimates, confidence intervals, and *P*-values are provided in Table S2 for the cross-ancestry analyses and Table S6 for the ancestry-specific analyses shown in this figure. Alt text: Twelve forest plots compare protein association estimates across European, East Asian, and African ancestry groups and the combined analysis. Squares show odds ratios, with horizontal 95% confidence intervals and a reference line at one.

Across the combined and separate East Asian outcome analyses, LRRC25, CASP8, ANXA4, and DNPH1 each had at least one nominally significant association (*P* < 0.05), with consistent effect directions between BCAC and BBJ (**Figure S2**).

Evidence of cross-ancestry heterogeneity was observed only for DNPH1 (Cochran’s Q test *P* < 0.05/12). Higher genetically predicted levels of DNPH1 were associated with higher odds of breast cancer in European ancestry (OR = 1.10, 95% confidence interval [CI]: 1.06 to 1.14, *P* = 4.10 × 10^-7^), showed a nominally inverse association in East Asian ancestry (OR = 0.80, 95% CI: 0.67 to 0.96, *P* = 1.63 × 10^-2^), and showed no evidence of association in African ancestry (OR = 0.98, 95% CI: 0.82 to 1.18, *P* = 0.85). When we repeated the East Asian analysis using *cis*-pQTL effect estimates derived from UKB-PPP East Asian-ancestry participants, the inverse association remained nominally significant and directionally consistent (IVW OR = 0.82, 95% CI: 0.70 to 0.97, *P* = 1.78 × 10^-2^).

### Associations with breast cancer intrinsic-like subtypes

We next evaluated the 12 candidate proteins for association with five intrinsic-like breast cancer subtypes in European-ancestry participants (**Figure 4**; **Table S7).** After Bonferroni correction (*P* < 0.05/12), 11 proteins (all except GCLM) were associated with at least one subtype, and all significant associations were directionally consistent with the overall breast cancer estimates. Significant associations were most frequent for luminal A-like disease (nine proteins), followed by luminal B-like and TNBC (four each), and HER2-enriched-like disease (one); none reached this threshold for luminal B/HER2-negative-like disease. Differences in statistical significance alone do not establish subtype specificity, particularly given unequal subtype sample sizes.

Three proteins showed statistically significant heterogeneity across subtypes after accounting for shared controls (Cochran’s Q test *P <* 0.05/12): CASP8 and RALB had their strongest associations with TNBC, whereas USP28 showed its strongest association with luminal A-like disease (**Figure 4**; **Table S7).**

**Figure 4.**
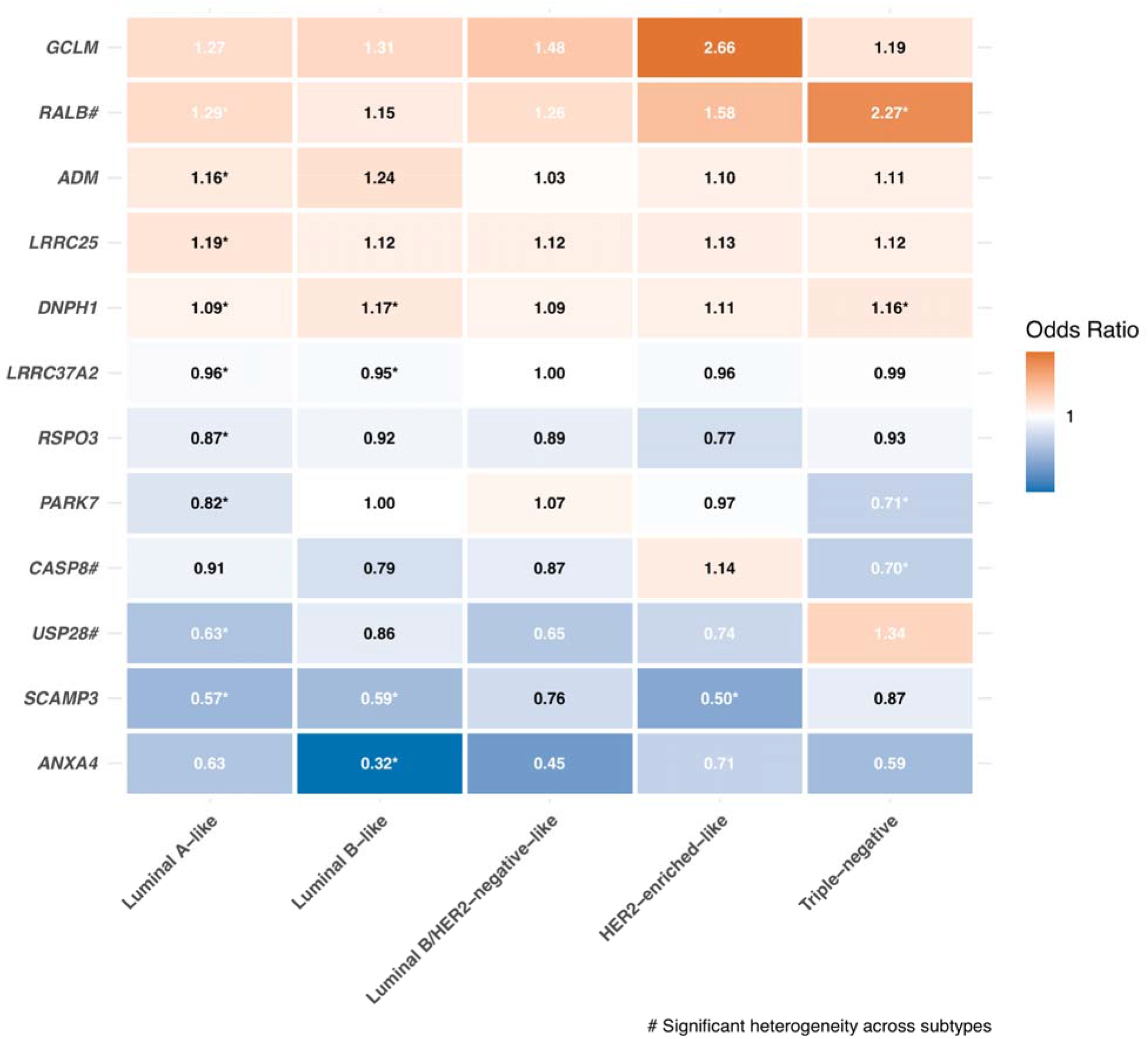
Heatmap of MR estimates for 12 candidate plasma proteins in relation to the risk of five intrinsic-like breast cancer subtypes in European-ancestry participants. Each tile represents the association between a given protein’s genetically predicted plasma level (y-axis) and a specific intrinsic-like breast cancer subtype (x-axis), derived from European-ancestry GWAS data. Tile shading indicates the direction and magnitude of the odds ratio (OR); the value within each tile is the OR point estimate. ORs above 1 indicate higher odds, and ORs below 1 indicate lower odds. Asterisks (*) indicate associations surviving Bonferroni correction across 12 proteins within each subtype (*P* < 0.05/12). A hash symbol (#) next to the protein name indicates significant heterogeneity of effects across subtypes by the covariance-adjusted heterogeneity test accounting for shared controls, with Bonferroni correction for 12 proteins (*P* < 0.05/12). Apparent subtype-specific patterns for proteins without significant heterogeneity should be interpreted cautiously, given the smaller sample sizes available for several non-luminal A subtype analyses. Alt text: Heatmap of odds ratios for 12 proteins across five breast cancer subtypes, with values printed in each cell. Asterisks identify associations meeting the corrected significance threshold. Three proteins have significant heterogeneity across subtypes.

### Reverse-direction Mendelian Randomization

We found no evidence that genetically proxied breast cancer liability influenced plasma levels of any of the 12 candidate proteins (all *P* > 0.10) (**Table S8).**

### Multi-layer Validation

**Table 2** summarizes the complementary evidence for the 12 candidate proteins across colocalization, external European (deCODE) and East Asian (JCTF) pQTL replication, and phenotype-linked PGS validation in AoU.

#### Colocalization analysis

Colocalization analyses in European-ancestry participants were performed to evaluate whether a shared causal variant within the *cis*-regulatory region drives the associations with both plasma protein levels and breast cancer risk. At a PPH4 threshold of ≥ 0.80, six of the 12 candidate proteins showed strong evidence of colocalization: LRRC25, DNPH1, ANXA4, CASP8, PARK7, and LRRC37A2 (**Table 2**, **Table S9).** Among these, four loci (LRRC25, DNPH1, CASP8, and LRRC37A2) contained variant(s) with ≥80% posterior probability of being the shared causal variant, conditional on H4 (SNP.PP.H4 ≥ 0.80).

#### External replication in an independent European pQTL resource

We repeated MR using independently selected deCODE *cis*-pQTLs and the European-ancestry breast cancer GWAS used in the primary analysis^36^ (**Table 2**). Ten proteins were measured in deCODE, and five yielded eligible *cis*-pQTLs. Four showed nominally significant associations consistent with the primary European-ancestry estimates: RALB and GCLM were associated with higher breast cancer odds, and PARK7 and RSPO3 were associated with lower odds (*P* < 0.05).

Genetically predicted ANXA4 levels showed a significant association with higher odds of breast cancer in deCODE (OR = 1.15, 95% CI: 1.11 to 1.19, *P* = 3.71 × 10^-15^), directionally opposite to the inverse association obtained using UKB-PPP European-ancestry pQTLs (OR = 0.56, 95% CI: 0.44 to 0.73, *P* = 1.15 × 10^-5^).

#### External replication in an independent East Asian pQTL resource

We repeated MR using independently selected JCTF *cis*-pQTLs and the East Asian-ancestry breast cancer GWAS used in the primary analysis (**Table 2**). All 12 candidates were measured in JCTF, and six yielded eligible instruments. LRRC25 and SCAMP3 showed nominally significant associations consistent with the primary East Asian estimates (*P* < 0.05). For DNPH1, the single eligible instrument yielded a directionally consistent, though non-significant, inverse association (OR = 0.93, 95% CI: 0.59 to 1.47, *P* = 0.75).

#### Phenotype-linked genetic validation in the All of Us cohort

We constructed per-protein PGS and tested their associations with observed breast cancer status in the independent, multi-ancestry AoU cohort (9,250 cases and 214,857 controls; **Table 2**; **Table S10**).

In the pooled multi-ancestry analysis, four proteins showed nominally significant associations in the same direction as the primary cross-ancestry meta-analysis estimates: higher PGSs for LRRC37A2, SCAMP3, and ANXA4 were associated with lower odds of breast cancer, whereas a higher PGS for LRRC25 was associated with higher odds (*P* < 0.05).

Ancestry-stratified analyses yielded one additional directionally concordant finding: the protective association of USP28 was supported in the East Asian stratum. ADM, which showed a null association in our primary African-specific MR, demonstrated a nominally significant inverse association in the AoU African stratum.

### Summary across validation layers

The 12 candidate proteins differed in the extent of complementary support across validation analyses (**Table 2**). The strongest overall support was observed for LRRC25, PARK7, and LRRC37A2, each corroborated by the primary MR analysis, strong colocalization, and at least one external replication or phenotype-linked validation result. CASP8 and DNPH1 showed strong primary MR and colocalization corroboration, although external support was limited for CASP8 and interpretation of DNPH1 is complicated by cross-ancestry heterogeneity. RALB, GCLM, SCAMP3, RSPO3 and USP28 were supported by the primary MR analysis together with at least one additional validation layer, but lacked strong colocalization evidence. ADM and ANXA4 should be considered more provisional: ADM had limited support across validation analyses, whereas ANXA4 showed platform-dependent directional inconsistency.

## Discussion

This proteome-wide MR study identifies 12 plasma proteins associated with breast cancer risk and shows that support for these associations varies across ancestry-specific, subtype-specific, and validation analyses. By combining cross-ancestry MR with colocalization, external proteomic replication, and phenotype-linked genetic validation, we prioritized a subset of candidates with stronger cumulative evidence for involvement in breast cancer susceptibility. Collectively, these findings nominate genetically prioritized plasma proteins for mechanistic investigation and translational follow-up.

Our findings both reinforce prior work and extend it. Among our 12 candidate proteins, eight had previously been linked to breast cancer risk by MR, all concordant in direction with the reported significant associations in those studies^23,25–29^. Four additional candidates emerged from our analyses. Among these, GCLM, ANXA4, and USP28 had been examined in prior MR: GCLM and USP28 yielded directionally consistent estimates that did not meet the original studies’ corrected significance criteria, while ANXA4 showed a supplementary MR signal with weak integrated support. Our study extends prior work through cross-ancestry and subtype comparisons and complementary evaluation of the candidate proteins. Differences in protein coverage, genetic instruments, outcome datasets, and selection criteria may explain variation in findings across studies. Colocalization supported a shared association signal for six proteins, although it does not by itself establish that these proteins mediate the association with breast cancer. Causal interpretation requires greater caution for proteins without strong colocalization evidence^47^.

Several prioritized proteins have plausible biological links to breast cancer. For example, CASP8, a key initiator of extrinsic apoptosis, has long been implicated in breast cancer susceptibility^48,49^. Transcriptome-wide association studies have linked higher genetically predicted CASP8 expression to reduced breast cancer risk in European and Asian populations, with nominal replication in African analyses^50–52^. ADM, SCAMP3, and USP28 also have biologically plausible roles through angiogenesis, membrane trafficking, and protein stability, respectively, and have been implicated in breast cancer cell proliferation or survival^53–56^. While these observations do not establish mechanism, they support the biological plausibility of the prioritized proteins.

Previous MR studies reported associations with other cancers for ten of our 12 candidate proteins (**Table S11**). These cross-cancer associations may reflect involvement in tumorigenic programs shared across cancer types, although their implications for biomarker specificity require direct evaluation. Among the proteins with the strongest cumulative support, PARK7 (DJ-1) is widely expressed^57^ and is implicated in autosomal recessive early-onset parkinsonism^58^; it has also been linked to other cancers, with upregulation in oral squamous cell carcinoma tissue and reduced proliferation and invasion upon knockdown in oral cancer cells^59^. LRRC25 is expressed in myeloid cells, where mouse tumor models indicate an immunosuppressive role^60^.

LRRC37A2 shows testis-enriched RNA expression^61^ and has also been associated with Parkinson’s disease risk^62,63^. Associations with other conditions do not preclude a role in breast cancer etiology, provided the pathways these proteins act through are active in breast tissue, through local and/or systemic effects. How these tissue-level functions relate to genetically predicted circulating levels, however, remains unresolved.

A notable finding was the directionally heterogeneous association of DNPH1 across ancestries. This pattern could reflect true biological heterogeneity, but it could also arise from imperfect transferability of European-derived instruments to non-European populations, given ancestry differences in LD structure, allele frequency, and regulatory architecture. Complementary analyses across East Asian UKB-PPP and JCTF pQTL datasets suggested that the directional difference is not fully explained by European-derived effect estimates alone. Biologically, DNPH1 has been linked to sensitivity to poly(ADP-ribose) polymerase (PARP) inhibition in BRCA1/2-deficient cells^64^, and population differences in BRCA founder mutations^65^ suggest a possible source of variation in its biological relevance, although this explanation remains speculative. More broadly, this result illustrates both the value and the current limitations of cross-ancestry proteogenomic inference, motivating the development of larger ancestrally diverse pQTL resources.

Subtype-stratified analyses further suggested etiological heterogeneity for CASP8, RALB and USP28. Higher genetically predicted CASP8 levels were most strongly associated with reduced TNBC risk, consistent with experimental evidence linking caspase-8 activation to stress-induced apoptosis in TNBC cells^66^ and supporting further investigation of apoptotic pathways in TNBC susceptibility. RALB, a small GTPase implicated in TNBC cell survival^67^, similarly showed the strongest risk-increasing association with TNBC, supporting further investigation of its role in this subtype. USP28 showed its most pronounced protective association with the luminal A-like subtype alongside a positive, non-significant association in TNBC. Given evidence that USP28 can stabilize oncogenic substrates and promote breast cancer cell survival^56,68^, these results suggest that its net effect may depend on tumor context.

Proteins with broader, subtype-spanning associations may act through susceptibility pathways shared across breast cancer subtypes. These findings underscore the molecular heterogeneity of breast cancer susceptibility and highlight the value of subtype-resolved proteomic evidence for guiding precision prevention and therapeutic prioritization.

Our external validations using independent pQTL resources provided additional support for a subset of findings. In deCODE, ANXA4 showed a directionally opposite association relative to UKB-PPP, consistent with previous reports that antibody-and aptamer-based assays can capture distinct molecular features and yield discordant genetically predicted disease associations^69,70^. Given the multiple isoforms and conformation-dependent membrane states of annexins^71^, platform-dependent capture of different molecular forms is a plausible explanation, although the direction of effect for ANXA4 remains uncertain. Replication of LRRC25 and SCAMP3 in the considerably smaller JCTF resource provides further, though limited, support using East Asian-derived genetic instruments. PGS analyses in AoU replicated LRRC37A2, SCAMP3, ANXA4, and LRRC25 in the pooled analysis, providing additional support in an outcome cohort independent of the primary GWAS. Because these analyses do not involve direct protein measurement, they complement rather than replace MR and colocalization evidence.

Our findings carry implications for mechanistic research and translational prioritization. Together, the forward and reverse MR findings are more consistent with a role for these proteins in susceptibility-related pathways than with an exclusively disease-reactive interpretation. While these targets remain provisional, preclinical studies have demonstrated covalent targeting of GCLM, the modifier subunit of glutamate-cysteine ligase, and inhibition of DNPH1 in cancer models^64,72^; LRRC25 has been proposed as a potential cancer immunotherapy checkpoint target and peptides derived from LRRC37A2-containing fusion transcripts have been investigated as potential breast cancer vaccine candidates^60,73^. Three priorities would advance this field: generation of large-scale ancestrally diverse pQTL resources to enable more robust non-European instrument discovery; prospective studies assessing protein-based risk prediction and early detection; and functional validation to establish the mechanistic roles of candidate proteins in breast cancer tumorigenesis.

Several limitations warrant consideration. Our primary instruments were derived from European-ancestry participants because comparably large non-European pQTL resources are not yet available. The cross-ancestry meta-analysis is therefore not a symmetric multi-ancestry analysis; it is more accurately characterized as a cross-ancestry outcome evaluation using predominantly European-derived instruments, with associations substantially driven by European data. We note that UKB-PPP reported strong concordance of *cis*-pQTL effect sizes across ancestry groups, providing empirical justification for our primary approach^17^. Ancestry classifications were also source-defined and not uniformly harmonized across datasets, and broad ancestry labels obscure within-group diversity, which could not be assessed from the available pooled summary statistics. Subtype-stratified analyses were restricted to European-ancestry data, and the smaller case counts for several subtypes limit statistical power for detecting subtype-specific associations. The shared-control adjustment used a covariance approximation because covariance matrices from the fitted subtype GWAS model were unavailable. We did not directly assess whether genetic effects on protein levels vary with age. If they do, the *cis*-pQTL weights estimated in the exposure sample may not fully transport to the outcome populations, potentially biasing the MR estimates. Sex-combined pQTL estimates were used in the primary analyses to maximize exposure sample size and instrument availability. The high concordance between the female-only and sex-combined estimates supports the robustness of the evaluated associations, although the smaller female-only sample and incomplete instrument coverage limit power to assess sex-specific effects. Restricting instruments to *cis*-pQTLs mitigates concerns about horizontal pleiotropy, but, even with our alternative MR analyses, does not exclude effects on breast cancer through pathways independent of the target protein. Finally, MR estimates reflect lifelong genetically predicted protein levels and may not predict how pharmacological intervention would act at a specific age or disease stage.

This cross-ancestry proteome-wide MR study identified 12 plasma proteins associated with breast cancer risk, including ancestry-and subtype-differentiated signals, and prioritized a subset with greater cumulative support from colocalization, external MR, and protein PGS analyses. These findings provide a genetically informed basis for mechanistic and translational follow-up, and motivate the larger, ancestrally diverse pQTL resources needed for equitable target and biomarker discovery.

## Acknowledgements

We gratefully acknowledge the researchers and consortia who generated and made publicly available the GWAS summary statistics used in this study. We thank the All of Us Research Program participants and staff, the participants of all contributing GWAS cohorts, and the investigators whose efforts made these datasets accessible for secondary analysis.

ChatGPT (OpenAI) was used to assist with language editing and grammatical corrections in the manuscript and to help develop portions of the code used for data analyses. The authors directed the use of the tool, reviewed its output, and performed and reviewed all analyses. The authors take full responsibility for the accuracy and integrity of the analyses and the manuscript.

The contributions of the NIH authors are considered Works of the United States Government. The findings and conclusions presented in this paper are those of the authors and do not necessarily reflect the views of the NIH or the U.S. Department of Health and Human Services.

The funder had no role in the design of the study; the collection, analysis, or interpretation of the data; or the writing of the manuscript and decision to submit it for publication.

## Data Availability

Summary-level GWAS statistics for plasma proteomics data are available through the UK Biobank (https://www.synapse.org/Synapse:syn51364943/), deCODE genetics (https://www.decode.com/), and the Japan COVID-19 Task Force (https://humandbs.dbcls.jp/files/hum0343/hum0343.v3.qtl.v1.zip). Summary-level GWAS statistics for breast cancer are available through https://www.ccge.medschl.cam.ac.uk/breast-cancer-association-consortium-bcac (European-ancestry), https://www.ccge.medschl.cam.ac.uk/breast-cancer-association-consortium-bcac/data-data-access/summary-results/gwas-summary-results, https://pheweb.jp/pheno/BrC (East Asian-ancestry), and https://www.ebi.ac.uk/gwas/studies/GCST90296719 (African-ancestry). Individual-level phenotype data and whole-genome sequencing WGS data from the All of Us are available to registered researchers through the All of Us Researcher Workbench (https://workbench.researchallofus.org/). Custom code used for data harmonization, Mendelian randomization analyses, colocalization, and validation analyses is publicly available at https://github.com/NCI-CGR/mr_breast_cancer.

