## Supplementary Methods for "Cross-ancestry proteome-wide Mendelian randomization of plasma proteins and breast cancer susceptibility"

### Supplementary Material

#### Supplementary Methods

#### S1 Shared-control covariance in subtype-specific MR

##### S1.1 Setting and first-order inference

For a given exposure, let $\theta_s$ denote its Mendelian randomization (MR) association with disease subtype $s$, expressed as a log odds ratio per unit increase in the genetically predicted exposure. Let $K \geq 2$ be the number of mutually exclusive disease subtypes compared against a shared control group. The target null hypothesis is

$$H_0 : \theta_1 = \cdots = \theta_K. \tag{S1}$$

Indices $s, t \in \{1, \ldots, K\}$ label subtypes. Let $\mathcal{J}_s$ be the retained SNP instrument set for subtype $s$, $J_s = |\mathcal{J}_s|$ its size, and $\mathcal{J} = \bigcup_{s=1}^{K} \mathcal{J}_s$ the union, of size $J$. Indices $j$ and $\ell$ label SNPs in this union. Write $g_j = \widehat{\gamma}_j$ for the estimated SNP-exposure association, where $\gamma_j$ is the population association per copy of the effect allele. Let $b_{js}$ be the estimated SNP-subtype log odds ratio and $\sigma_{js} = \mathrm{SE}(b_{js})$ its reported standard error. In particular, $\sigma_{j1}, \ldots, \sigma_{jK}$ are the standard errors for SNP $j$ in subtypes $1, \ldots, K$, respectively. All associations are harmonized to the same effect allele.

The general notation allows instrument sets to differ when outcome availability differs. When every retained SNP is available for every subtype, $\mathcal{J}_s = \mathcal{J}$ and $J_s = J$ for all $s$; no missing-instrument adjustment is then needed. Define the $J \times 1$ vector $g = (g_j)_{j \in \mathcal{J}}$ and the collection of observed outcome standard errors $\mathcal{S} = \{\sigma_{js} : s = 1, \ldots, K,\ j \in \mathcal{J}_s\}$. The derivation conditions on the retained instrument sets, $g$, and $\mathcal{S}$. It therefore follows first-order inverse-variance weighted (IVW) inference [2]: uncertainty in $g_j$ is omitted, and no second-order ratio-variance term is added. This is an inferential approximation, not an assertion that exposure associations are measured without error. Throughout, $\mathrm{diag}(x_1, \ldots, x_r)$ denotes a diagonal matrix with those entries, and the superscript $\top$ denotes transpose.

We assume negligible residual linkage disequilibrium among retained instruments and negligible exposure-outcome sample overlap. For the sampling model, errors for different SNPs are treated as independent, whereas errors for the same SNP may be correlated across subtypes. Usual MR identification assumptions are required for causal interpretation; covariance adjustment alone does not address instrument invalidity.

##### S1.2 Covariance matrix for the SNP-subtype associations

Let $n_s$ be the number of cases for subtype $s$ and $n_0$ the common number of controls. Let $\rho_{st}$ denote the working correlation between sampling errors in $b_{js}$ and $b_{jt}$, assumed constant across SNPs. With disjoint subtype case groups and completely shared controls, the sample-overlap formula of Lin and Sullivan [1] reduces, under a small-effect/null approximation, to

$$\rho_{st} \approx \frac{1/n_0}{\sqrt{(1/n_s + 1/n_0)(1/n_t + 1/n_0)}} = \sqrt{\frac{n_s n_t}{(n_s + n_0)(n_t + n_0)}}, \qquad s \neq t. \tag{S2}$$

One way to see this reduction is to write the approximate sampling error of a SNP effect as a subtype-case contribution minus a common control contribution. Let $G_j$ denote the effect-allele dosage at SNP $j$ for a sampled individual, and define $v_j = \mathrm{Var}(G_j) > 0$ as its population variance, approximated as common across the case and control groups. The marginal variance of $b_{js}$ is then approximately $v_j^{-1}(1/n_s + 1/n_0)$, and the covariance between $b_{js}$ and $b_{jt}$ is approximately $v_j^{-1}/n_0$. The genotype factor cancels in equation (S2); $v_j$ is used to explain the approximation and is not a required pipeline input. Strong genetic or covariate effects can affect its accuracy.

Incomplete subtype classification arises when one or more tumor characteristics needed to assign a subtype are unavailable. For example, the Breast Cancer Association Consortium (BCAC) analysis by Zhang et al. used a two-stage polytomous logistic regression model with an expectation-maximization (EM) algorithm to incorporate cases with missing estrogen receptor, progesterone receptor, HER2, or tumor-grade information [3]. Its published Supplementary Note describes the estimation of information-equivalent effective case counts and their summarization by the median across SNPs. When fitted covariance matrices are unavailable, effective case counts obtained by this method can be substituted for $n_s$ in equation (S2) to obtain a working shared-control correlation. These are effective case counts, not effective total case-control sample sizes.

When available, the SNP-specific covariance matrix of the final subtype log odds ratio estimates is preferred and can be propagated directly through the IVW estimator below. It can account for dependence introduced by joint estimation, including EM estimation, even when sample sizes are constant. This is particularly useful when the analyzed sample sizes or their overlap across subtypes vary substantially across SNPs, because a single count-based correlation matrix may then be inadequate. Under the count-based approximation, the actual SNP-level standard errors continue to determine the marginal variances.

Set $\rho_{ss} = 1$ and define the $K \times K$ outcome-error correlation matrix $R$ by $R_{st} = \rho_{st}$. For SNP $j$ observed in all subtypes, let $b_j = (b_{j1}, \ldots, b_{jK})^\top$ be its $K \times 1$ outcome-association vector, and let $S_j = \mathrm{diag}(\sigma_{j1}, \ldots, \sigma_{jK})$ be the $K \times K$ diagonal matrix of its reported standard errors. The $K \times K$ working sampling covariance of $b_j$ is

$$C_j = S_j R S_j, \qquad (C_j)_{st} = \rho_{st}\sigma_{js}\sigma_{jt}. \tag{S3}$$

Thus, the actual GWAS standard errors determine the marginal variances; sample sizes determine only the working correlations. For a SNP with missing outcome estimates, we form the covariance on its available subtypes and embed it in a $K \times K$ matrix with unobserved rows and columns set to zero.

##### S1.3 IVW estimator covariance matrix

For each subtype, the usual unpenalized first-order IVW estimator minimizes

$$\sum_{j \in \mathcal{J}_s} \frac{(b_{js} - g_j\theta_s)^2}{\sigma_{js}^2}. \tag{S4}$$

Differentiation with respect to $\theta_s$ gives

$$D_s = \sum_{j \in \mathcal{J}_s} \frac{g_j^2}{\sigma_{js}^2}, \qquad \widehat{\theta}_s = \frac{\sum_{j \in \mathcal{J}_s} g_j b_{js}/\sigma_{js}^2}{D_s} = \sum_{j \in \mathcal{J}_s} a_{js} b_{js}, \qquad a_{js} = \frac{g_j}{\sigma_{js}^2 D_s}. \tag{S5}$$

Here $D_s$ is the first-order information for subtype $s$, and $a_{js}$ is the linear coefficient multiplying $b_{js}$ in its IVW estimate. We require $D_s > 0$ and set $a_{js} = 0$ for unavailable SNPs. Conditional on the exposure estimates and weights, this estimator is linear in the outcome estimates. Let $V^{(0)}$ be the $K \times K$ covariance matrix of the subtype MR estimates under the sampling-only working model. Its $(s,t)$ entry is

$$\begin{aligned} V_{st}^{(0)} &:= \operatorname{Cov}(\widehat{\theta}_s, \widehat{\theta}_t \mid g, \mathcal{S}) \\ &= \sum_{j \in \mathcal{J}_s} \sum_{\ell \in \mathcal{J}_t} a_{js} a_{\ell t} \operatorname{Cov}(b_{js}, b_{\ell t} \mid g, \mathcal{S}) \\ &= \sum_{j \in \mathcal{J}_s \cap \mathcal{J}_t} a_{js} a_{jt} (C_j)_{st} \\ &= \frac{\rho_{st}}{D_s D_t} \sum_{j \in \mathcal{J}_s \cap \mathcal{J}_t} \frac{g_j^2}{\sigma_{js} \sigma_{jt}}. \end{aligned} \tag{S6}$$

The last equality uses the SNP-invariant working correlation in equation (S2). The preceding covariance-propagation identity also applies to SNP-specific $C_j$ supplied by the original outcome model. In particular,

$$V_{ss}^{(0)} = \sum_{j \in \mathcal{J}_s} a_{js}^2 \sigma_{js}^2 = D_s^{-1}. \tag{S7}$$

For a matrix representation, define the $K \times K$ diagonal coefficient matrix $A_j = \operatorname{diag}(a_{j1}, \ldots, a_{jK})$. Then

$$V^{(0)} = \sum_{j \in \mathcal{J}} A_j C_j A_j^{\top}. \tag{S8}$$

Unavailable components have zero coefficient; only observed principal submatrices contribute. Each summand is positive semidefinite. Equations (S6) and (S8) provide the extension from association-level shared-control covariance to MR-level covariance. The linear propagation is exact conditional on the supplied $C_j$ and fixed weights; the sample-size construction of $C_j$ remains approximate.

The outcome-error correlation generally differs from the MR-estimate correlation. Writing $\operatorname{Corr}_0$ for correlation under the sampling-only working model,

$$\operatorname{Corr}_0(\widehat{\theta}_s, \widehat{\theta}_t) = \rho_{st} \frac{\displaystyle\sum_{j \in \mathcal{J}_s \cap \mathcal{J}_t} g_j^2 / (\sigma_{js} \sigma_{jt})}{\sqrt{D_s D_t}}. \tag{S9}$$

The factor multiplying $\rho_{st}$ is the normalized inner product of two vectors with entries $g_j/\sigma_{js}$, padded with zeros for unavailable SNPs. It lies between zero and one. It equals one when these vectors are proportional, including the case in which both subtypes use the same single instrument. Therefore, directly combining $R$ with final MR standard errors generally omits the effect of subtype-specific weights and instrument availability.

When all subtypes use the same single SNP $j$, with $g_j \neq 0$, the estimator reduces to the Wald ratio. Its first-order variance and covariance are

$$\widehat{\theta}_s = \frac{b_{js}}{g_j}, \qquad V_{ss}^{(0)} = \frac{\sigma_{js}^2}{g_j^2}, \qquad V_{st}^{(0)} = \frac{(C_j)_{st}}{g_j^2} = \frac{\rho_{st} \sigma_{js} \sigma_{jt}}{g_j^2}. \tag{S10}$$

#### S1.4 Compatibility with multiplicative random-effects IVW

For compatibility with `mr_ivw` using simple weights and its default model, the marginal variance specification is fixed-effect variance for two or three instruments and multiplicative random-effects variance for four or more; a single instrument uses the first-order Wald ratio [4]. The point estimator in equation (S5) is unchanged by a subtype-specific multiplicative variance factor. Define the within-subtype residual statistic and standard-error inflation factor by

$$Q_{\mathrm{res},s} = \sum_{j \in \mathcal{J}_s} \frac{(b_{js} - g_j \widehat{\theta}_s)^2}{\sigma_{js}^2}, \qquad h_s = \begin{cases} 1, & J_s \le 3, \\ \max\{1, \sqrt{Q_{\mathrm{res},s}/(J_s - 1)}\}, & J_s \ge 4. \end{cases} \tag{S11}$$

The default marginal MR variance is $h_s^2/D_s$. This instrument-level residual statistic is distinct from the test of equality across subtypes below. First-order weights and multiplicative dispersion address different issues; retaining $h_s$ does not introduce exposure-effect uncertainty.

Separate univariate dispersion estimates do not determine cross-subtype covariance of excess residual variation. We therefore specify a joint working model. For each observed SNP-subtype pair, write

$$b_{js} = g_j \theta_s + e_{js} + \eta_{js}, \qquad \mathrm{Cov}(e_j) = C_j, \tag{S12}$$

where $e_{js}$ is the GWAS sampling error and $\eta_{js}$ is an additional instrument residual for SNP $j$ and subtype $s$. Their $K \times 1$ vectors are $e_j = (e_{j1}, \ldots, e_{jK})^\mathsf{T}$ and $\eta_j = (\eta_{j1}, \ldots, \eta_{jK})^\mathsf{T}$. Both have conditional mean zero given $g$ and $\mathcal{S}$, and the two components are independent. For missing subtype entries, the vectors are padded with zeros solely for matrix notation. For the main working analysis, assume that the observed $\eta_{js}$ are independent across subtypes and across SNPs, with

$$\mathrm{Var}(\eta_{js}) = (h_s^2 - 1)\sigma_{js}^2. \tag{S13}$$

Treating the estimated $h_s$ as plug-in quantities and propagating both components gives

$$V^{\mathrm{main}} = V^{(0)} + \mathrm{diag}\left\{(h_s^2 - 1)V_{ss}^{(0)} : s = 1, \ldots, K\right\}. \tag{S14}$$

This choice retains the shared-control sampling covariances and reproduces the original default marginal variances. Its substantive additional assumption is that excess instrument residual variation is uncorrelated between subtypes.

#### S1.5 Omnibus test of equality across subtypes

Collect the estimates in the $K \times 1$ vector $\widehat{\theta} = (\widehat{\theta}_1, \ldots, \widehat{\theta}_K)^\mathsf{T}$. Let $\mathbf{1}_r$ denote the $r \times 1$ vector of ones and $I_r$ the $r \times r$ identity matrix. Define the full-row-rank $(K-1) \times K$ contrast matrix

$$L = \begin{bmatrix} -\mathbf{1}_{K-1} & I_{K-1} \end{bmatrix}, \qquad d = L\widehat{\theta}, \qquad W = LVL^\mathsf{T}. \tag{S15}$$

Here $V = V^{\mathrm{main}}$ is the $K \times K$ MR covariance matrix in equation (S14), $d$ is the $(K-1) \times 1$ vector of estimated differences from subtype 1, and $W$ is its $(K-1) \times (K-1)$ covariance matrix. The omnibus statistic is

$$Q_{\mathrm{sub}} = d^\mathsf{T} W^{-1} d. \tag{S16}$$

Under the equality null, asymptotic normality, and a consistent estimator of the covariance matrix, the reference distribution is $\chi^2_{K-1}$ for $K$ subtypes. Because correlations and dispersion factors are approximated or estimated, the operational calibration is approximate, particularly with few instruments. The statistic is invariant to the reference subtype and to any nonsingular change of contrast basis. A Cholesky solve is used in implementation; singular or ill-conditioned contrast covariances trigger an error rather than automatic matrix repair.

When $V$ is positive definite, let $\widehat{\mu}$ be the generalized least-squares estimate of the common subtype effect under $H_0$. With $\mathbf{1} = \mathbf{1}_K$, the same statistic can be written as a generalized Cochran statistic:

$$\widehat{\mu} = \frac{\mathbf{1}^\top V^{-1}\widehat{\theta}}{\mathbf{1}^\top V^{-1}\mathbf{1}}, \qquad Q_{\text{sub}} = (\widehat{\theta} - \widehat{\mu}\mathbf{1})^\top V^{-1}(\widehat{\theta} - \widehat{\mu}\mathbf{1}). \tag{S17}$$

For two subtypes this reduces to

$$Q_{\text{sub}} = \frac{(\widehat{\theta}_s - \widehat{\theta}_t)^2}{V_{ss} + V_{tt} - 2V_{st}}. \tag{S18}$$

Thus, positive shared-control covariance reduces the variance of a difference when marginal variances are held fixed. The test assesses equality of subtype effects, not whether any individual subtype effect differs from zero. For a prespecified family of $M$ exposure comparisons, a Bonferroni significance threshold is $P < \alpha/M$, where $\alpha$ is the family-wise error level.

##### S1.6 Application to breast cancer subtypes and interpretation

For the present protein MR analysis, $K = 5$ breast cancer subtypes are compared using European-ancestry Breast Cancer Association Consortium (BCAC) summary statistics. The triple-negative outcome is restricted to BCAC alone. Effects are expressed per one standard deviation increase in genetically predicted protein abundance. Accordingly, the contrast matrix has dimension $4 \times 5$, and the reference distribution is $\chi^2_4$. With $M = 12$ prespecified protein comparisons and $\alpha = 0.05$, the significance threshold is $P < 0.05/12$.

For each protein, all retained SNP instruments are available across the five subtype outcomes, so $\mathcal{J}_1 = \cdots = \mathcal{J}_5 = \mathcal{J}$ and $J_s = J$. Thus, the intersections in equations (S6) and (S9) reduce to the common instrument set. The subtype-specific outcome standard errors, and therefore the IVW coefficients $a_{js}$, may still differ; a common instrument set alone does not make the MR-estimate correlation equal to the outcome-error correlation.

For this application, we use effective case counts supplied by the investigators of the source BCAC study (Table S12), together with $n_0 = 91{,}477$ shared controls. The method for estimating these counts is described in the published Supplementary Note [3]. The resulting working correlation represents the shared-control component of dependence. Covariance matrices from the fitted outcome model would also account for dependence arising from the joint use of partially observed tumor characteristics and the study's estimation procedure.

The analysis used effective case counts and SNP-level outcome standard errors to construct the covariance matrices in equation (S3). The MR covariance matrix $V^{\text{main}}$ in equation (S14) was then used in the omnibus test.

The original MR point estimates and default standard errors were reproduced from the retained, harmonized SNPs for all 60 protein-subtype combinations before interpreting the heterogeneity results. The method uses a common exposure scale, original SNP-level outcome standard errors, and the same instrument sets as the subtype analyses. It does not estimate covariance from the correlation of the 12 protein estimates or from genetic correlations between subtypes. Residual linkage disequilibrium, if material, requires cross-SNP covariance terms in equation (S6) and is outside this implementation.

Table S12: BCAC sample counts used in the shared-control covariance approximation.

| Subtype | Effective cases | Observed cases | Shared controls |
|---|---|---|---|
| Luminal A-like | 45,253 | 27,481 | 91,477 |
| Luminal B/HER2-negative-like | 6,350 | 6,799 | 91,477 |
| Luminal B-like | 6,427 | 6,502 | 91,477 |
| HER2-enriched-like | 2,884 | 2,793 | 91,477 |
| Triple-negative | 8,602 | 7,174 | 91,477 |

The effective case counts were supplied by the investigators of the source BCAC study and were not published in the original article or its supplementary materials. The published Supplementary Note describes their estimation method [3]. Effective counts need not exceed observed counts. The common control group is counted once, although its size appears in each row. No CIMBA participants enter this specification. Observed case counts are provided for context; the covariance calculation uses effective case counts.

The count-based covariance approximates shared-control dependence and does not reconstruct the covariance of the fitted EM estimator. The test also assumes that excess instrument residual variation is uncorrelated between subtypes, as specified in Section S1.4. Dispersion adjustment does not correct bias from directional pleiotropy.

Table S13 reports the corresponding heterogeneity $P$-values for the 12 proteins. CASP8, RALB, and USP28 met the significance threshold of $P < 0.05/12$.

Table S13: Covariance-adjusted tests of heterogeneity across breast cancer subtypes.

| Protein | Heterogeneity $P$-value |
|---|---|
| GCLM | 0.3287 |
| RALB | $1.53 \times 10^{-3*}$ |
| ADM | 0.6178 |
| LRRC25 | 0.5548 |
| DNPH1 | 0.5379 |
| ANXA4 | 0.4565 |
| SCAMP3 | 0.0238 |
| USP28 | $2.40 \times 10^{-4*}$ |
| CASP8 | $8.96 \times 10^{-5*}$ |
| PARK7 | 0.00471 |
| RSPO3 | 0.8270 |
| LRRC37A2 | 0.0318 |

The tests use the effective case counts and 91,477 shared controls in Table S12, the covariance matrix $V^{\text{main}}$ in equation (S14), and the omnibus statistic in equation (S16). In this application, $K = 5$ and each test has $K - 1 = 4$ degrees of freedom. Asterisks denote $P < 0.05/12$. These values correspond to the shared-control-adjusted heterogeneity results in Table S7.

### S2 Sensitivity to the cross-ancestry meta-analysis model

#### S2.1 Methods

To assess sensitivity to the cross-ancestry meta-analysis model, we compared fixed-effects meta-analysis, random-effects meta-analysis with between-ancestry variance estimated by restricted maximum likelihood (REML), and the Han and Eskin RE2 association test for the 12 prioritized proteins. The comparison uses the same available ancestry-specific MR log odds ratios and standard errors, expressed per one standard deviation increase in genetically predicted protein level. Fixed-effects meta-analysis was the primary discovery analysis; the alternative models were sensitivity analyses.

Within each ancestry-specific analysis, MR estimates were obtained using first-order (`simple`) IVW weights. Under the `MendelianRandomization` package defaults, analyses with two or three instruments used a fixed-effect model, and those with four or more used a multiplicative random-effects model. Single-instrument analyses used the Wald ratio with its first-order standard error. The standard errors supplied to each cross-ancestry meta-analysis retained these settings, including multiplicative variance inflation when applicable. Instrument-level heterogeneity within an MR analysis and heterogeneity between ancestry-specific MR estimates were therefore addressed at separate stages. Exposure-effect uncertainty was not added to the first-order MR variance.

Fixed-effects meta-analysis assumes a common underlying association and combines ancestry-specific estimates using inverse-variance weights. The random-effects model allows underlying associations to vary across ancestries. Between-ancestry variance was estimated by REML and added to each estimate's sampling variance when constructing inverse-variance weights. The Han and Eskin RE2 method, implemented in METASOFT, tests the global null of no association and no heterogeneity, allowing both the mean association and between-ancestry variance to be estimated under the alternative [5, 6]. RE2 therefore evaluates evidence for association in the presence of potentially heterogeneous effects.

#### S2.2 Interpretation

With at most three ancestry-specific estimates per protein, between-ancestry variance can be estimated imprecisely. This limits interpretation of the fitted heterogeneity and random-effects summaries; it does not by itself determine the validity of an association test. Model comparisons should be interpreted alongside ancestry-specific effect directions and heterogeneity tests. The proteome-wide significance threshold is $P < 0.05/1{,}971$, consistent with the primary analysis.

Statistical support may depend on the cross-ancestry meta-analysis model. RE2 tests a global null of no association while allowing heterogeneous effects under the alternative; a significant result does not establish a common effect or consistent direction across ancestries. These analyses use the same underlying estimates and do not provide independent replication.

Numerical comparisons are reported separately in Table S4.
