## Supplementary material for "Cross-ancestry proteome-wide Mendelian randomization of plasma proteins and breast cancer susceptibility": Figure S

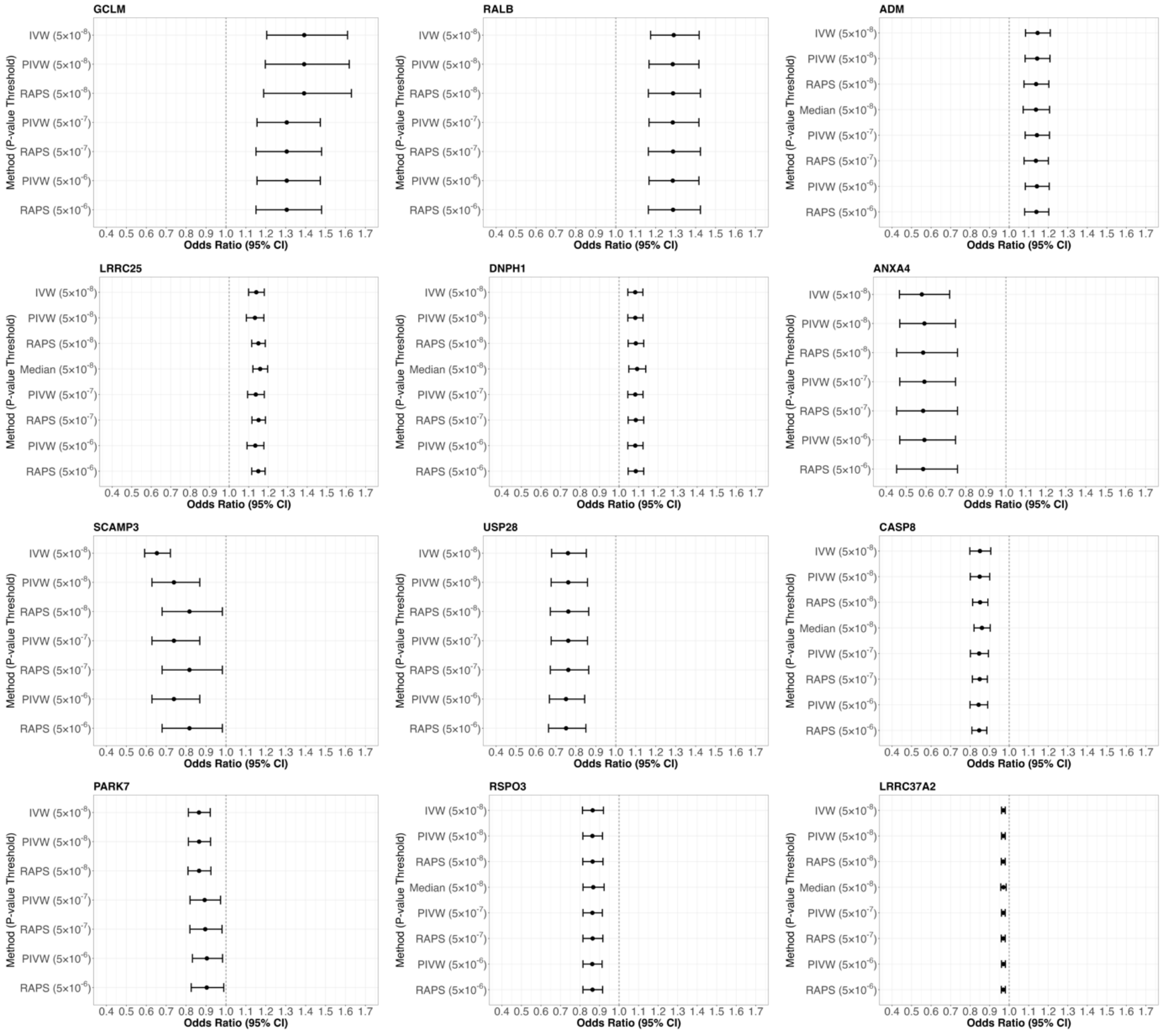
**Supplementary Material**

**Figure S1. Forest plots** **showing cross-ancestry associations between 12 candidate plasma proteins and overall breast cancer risk across sensitivity analyses.**

Each point represents the odds ratio (OR) per 1- standard deviation increase in genetically predicted protein levels, with horizontal lines indicating 95% confidence intervals. The x-axis is linear and capped at OR values of 0.4 and 1.7. Available primary and sensitivity estimates are shown for inverse-variance weighted (IVW), weighted median, penalized IVW (pIVW), and MR-Robust Adjusted Profile Score (MR-RAPS) analyses. The primary IVW and weighted-median analyses used instruments selected at *P* < 5 × 10^-8^. pIVW and MR-RAPS estimates are shown at *P* < 5 × 10^-8^, P < 5 × 10^-7^, and *P* < 5 × 10^-6^. Weighted-median estimates were unavailable for six proteins. Available ancestry-specific estimates were combined using fixed-effects meta-analysis. Full results are provided in Table S2.

Alt text: Twelve forest plots compare available estimates from inverse-variance weighting, weighted median, penalized inverse-variance weighting, and robust adjusted profile score methods under different instrument-selection thresholds. Association directions are consistent across available analyses, while confidence intervals and statistical support vary. Weighted-median results are unavailable for six proteins.

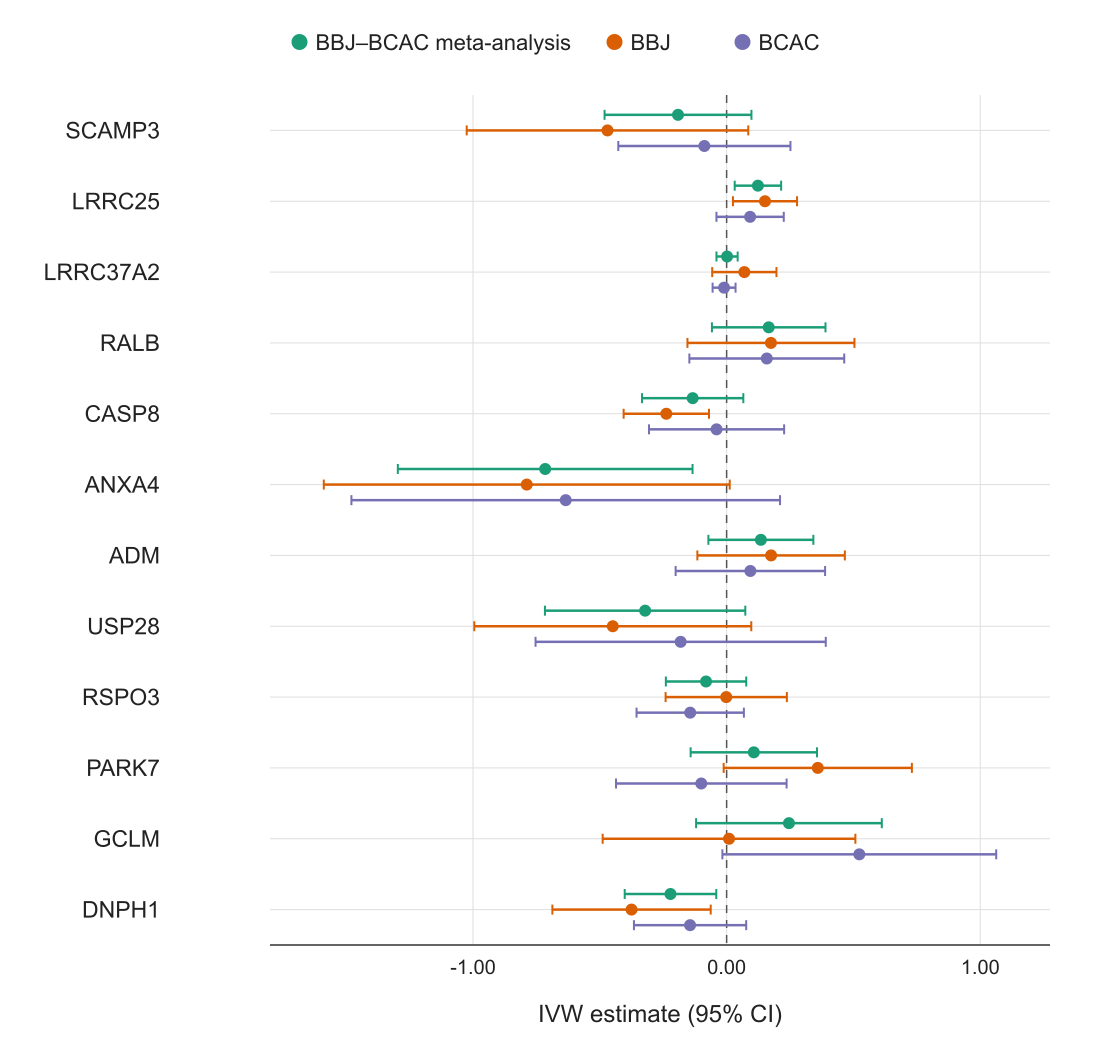
**Figure S2. East Asian-ancestry Mendelian randomization estimates for the 12 prioritized proteins by outcome source.**

Points show inverse-variance weighted (IVW) Mendelian randomization estimates using the combined BCAC and BBJ East Asian breast cancer GWAS, BBJ alone, and BCAC alone. Estimates are on the log-odds scale per one standard deviation increase in genetically predicted plasma protein level; horizontal lines indicate 95% confidence intervals. The dashed vertical line at zero indicates the null. Genetic instruments were derived from the UKB-PPP discovery pQTL dataset. BCAC, Breast Cancer Association Consortium; BBJ, BioBank Japan; UKB-PPP, UK Biobank Pharma Proteomics Project.

Alt text: Forest plot comparing East Asian breast cancer associations for 12 proteins using combined BCAC and BioBank Japan data and each source separately. Points show log-odds estimates with 95% confidence intervals; zero indicates no association.
